# Association of COVID-19 with risks of hospitalization and mortality from other disorders post-infection: A study of the UK Biobank

**DOI:** 10.1101/2022.03.23.22272811

**Authors:** Yong Xiang, Ruoyu Zhang, Jinghong Qiu, Hon-Cheong So

**Author notes:** Corresponding author* **Correspondence to: Hon-Cheong So**, MBBS, PhD. Lo Kwee-Seong Integrated Biomedical Sciences Building, The Chinese University of Hong Kong, Shatin, Hong Kong. Yong XIANG and Ruoyu ZHANG contributed equally to this manuscript.

## Abstract

**Objective:** To study whether COVID-19 infection may be associated with increased hospitalization and mortality from other diseases.

**Design:** Cohort study.

**Setting:** The UK Biobank.

**Participants:** All subjects in the UK Biobank with available hospitalization records and alive as of 31-Jan-2020 (*N*= 412,096; age 50-87).

**Main outcome measures:** We investigated associations of COVID-19 with hospitalization and mortality due to different diseases post-infection. We conducted a comprehensive survey on disorders from all systems (up to 135 disease categories). Multivariable Cox and Poisson regression was conducted controlling for main confounders. For sensitivity analysis, we also conducted separate analysis for new-onset and recurrent cases, and other analysis such as the prior event rate adjustment(PERR) approach to minimize effects of unmeasured confounders. We also performed association analyses stratified by vaccination status. Time-dependent effects on subsequent hospitalization and mortality were also tested.

**Results:** Compared to individuals with no known history of COVID-19, those with severe COVID-19 (requiring hospitalization) exhibited higher hazards of hospitalization and/or mortality due to multiple disorders (median follow-up=608 days), including disorders of respiratory, cardiovascular, neurological, gastrointestinal, genitourinary and musculoskeletal systems. Increased hazards of hospitalizations and/or mortality were also observed for injuries due to fractures, various infections and other non-specific symptoms. These results remained largely consistent after sensitivity analyses. Severe COVID-19 was also associated with increased all-cause mortality (HR=14.700, 95% CI: 13.835-15.619).

Mild (non-hospitalized) COVID-19 was associated with modestly increased risk of all-cause mortality (HR=1.237, 95% CI 1.037-1.476) and mortality from neurocognitive disorders, as well as hospital admission from a few disorders such as aspiration pneumonitis, musculoskeletal pain and other general signs/symptoms.

All-cause mortalities and hospitalizations from other disorders post-infection were generally higher in the pre-vaccination era. The deleterious effect of COVID-19 was observed to wane over time, with maximum HR in the initial phase.

**Conclusions:** In conclusion, this study revealed increased risk of hospitalization and mortality from a wide variety of pulmonary and extra-pulmonary diseases after COVID-19, especially for severe infections. Mild disease was also associated with increased all-cause mortality. Causality however cannot be established due to observational nature of the study. Further studies are required to replicate our findings.

## Introduction

There are more than 590 million^1^ documented cases of COVID-19 worldwide (as at 16-Aug-2022), and accumulating evidence have shown that the disease may be associated with various sequelae. For example, respiratory symptoms and disorders were reported frequently in the post-infection period, such as dyspnoea and cough^2^, lung function impairment^3^, and bronchiectasis^4^. Relatively less attention has been paid on other organ systems. For example, studies have reported that neurological manifestations could persist in patient with COVID-19^5-7^. Increased risks of cardiovascular diseases have also been observed^8^. Several other studies have also investigated the health consequences of COVID-19 across different systems^9-12^. Nevertheless, the full spectrum of sequalae from COVID-19 remains unclear, and there are still limited studies which have conducted a comprehensive analysis on potential sequelae from *all* body systems.

Given the very large number of people affected by COVID-19 worldwide, additional studies are still very much needed to delineate clearly the sequelae of COVID-19. There are also some limitations of existing studies. For example, some studies^11,12^ are of relatively small sample sizes, which limits the power to detect moderate associations. Another study^9^ examined the risks of a wide range of sequelae; however the study was restricted to veterans, who are predominantly males and the findings may not be readily generalizable to the general population^13^. In addition, very few works (except e.g. ref^14^) have studied how vaccination may change the risk of sequelae from COVID-19.

Here we leveraged a large prospective cohort of the UK biobank (UKBB) to identify associations of COVID-19 with hospitalization and mortality due to different diseases post-infection. We conducted a comprehensive survey on disorders from all systems, and a series of additional analysis, for example on the risk of hospitalization from new-onset and recurrent diseases, risk of sequelae across different follow-up periods, and how vaccination (before or after the infection) may alter the risk of sequelae.

## Methods

### UK Biobank sample

The UK Biobank (UKBB) is a large prospective cohort comprising ∼500,000 UK participants with continuous follow-up. The current age of subjects in this study ranged from 50 to 87 years. The details of the UKBB sample are described in ref^15^. As not every participant is linked to hospital inpatient records, our analysis was restricted to those subjects (sample size, *N*= 412,096) with available hospitalization records and alive as at 31-Jan-2020, the date for the first confirmed case of COVID-19 in UK. The current study was conducted under the project number 28732. The overall analytic flow is presented in Fig.1.

**Fig.1.**
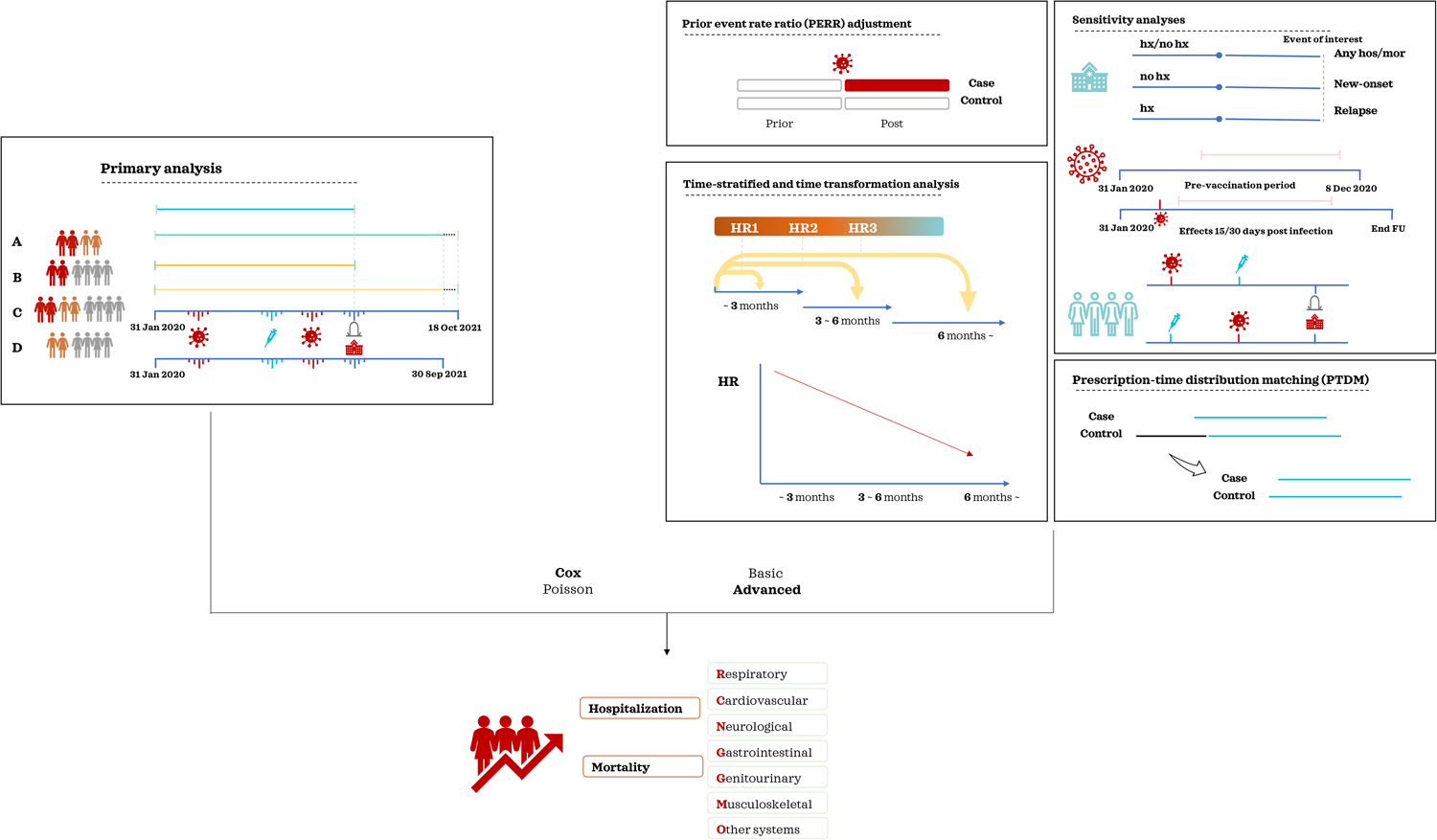
An overview of analysis flow. We investigated the associations between COVID-19 and subsequent risks of hospital admission/mortality in our primary analysis. For details please also refer to the Methods section in main text. We conducted analyses within infected patients (comparing outcomes of hospitalized vs non-hospitalized patients), and also compared hospitalized (severe) and non-hospitalized (mild) patients with subjects with no known history of infection. We conducted a comprehensive survey on disorders from all systems (up to 135 disease categories). Multivariable Cox and Poisson regression was conducted controlling for main confounders. For sensitivity analysis, we also conducted separate analysis for new-onset and recurrent cases, and other sensitivity analysis such as the prior event rate adjustment (PERR) approach to minimize effects of unmeasured confounders. We also performed associations analyses stratified by vaccination status, and evaluated the effects of infection restricted to the pre-vaccination period. Time-dependent effects on subsequent hospitalization and mortality were also tested.

### Study Outcomes

Our main study outcomes were hospitalization and mortality due to various disorders. Hospitalization records were extracted from the Hospital Episode Statistics (HES) data of UKBB. Detailed descriptions of HES can be found in https://biobank.ndph.ox.ac.uk/ukb/ukb/docs/HospitalEpisodeStatistics.pdf. The hospital inpatient ‘core’ and diagnoses datasets were updated to 30 Sep 2021 (accessed 21 Dec 2021). The diagnosis codes and corresponding dates were summarized based on each participant’s inpatient record. All the diagnosis codes were converted to 3-character ICD-10 codes. We followed the mapping strategy as described in https://biobank.ndph.ox.ac.uk/showcase/showcase/docs/first_occurrences_outcomes.pdf. The mortality records of UKBB contain the cause of death (ICD10-coded) and were linked to national death registries. Details can be found in https://biobank.ctsu.ox.ac.uk/crystal/crystal/docs/DeathLinkage.pdf. Mortality data were updated to 18 Oct 2021 (accessed 21 Dec 2021).

All ICD-10 codes were covered in analysing the sequelae of COVID-19 infection. We employed the latest version (2022.1) of Clinical Classifications Software Refined (CCSR)^16^ to classify ICD-10 codes to a smaller number of clinically meaningful categories of diagnoses. Totally 320 clinical categories covering 21 body systems were included as outcomes. Since the number of events may be very small for some outcomes, we only present the results if the number of events>=5 for both exposed and unexposed groups. Totally up to 135 disease categories were included after the filtering.

Only primary causes of admission/mortality were considered. All diagnosis given before the study start-date (i.e. time0; the date a subject entered the cohort study) were regarded as medical history of comorbidities; diagnoses given after time0 were treated as hospitalization/mortality due to ‘new-onset’ diseases. For better statistical power, we primarily present the results from *any* hospitalization or mortality from diseases, without consideration of past medical history. Nevertheless, we also performed additional stratified analysis on new-onset (subjects with no known prior history) and recurrent/relapsed diseases (subjects with known history of the disease).

### Covariates

We performed multivariable regression analysis with adjustment for covariates including basic demographics (age, sex, ethnicity), major risk factors of cardiovascular disorders (lipid and glucose levels, HbA1c), immunological disorders (autoimmune diseases, immunodeficiency, history of receiving immunosuppressants), major comorbidities (coronary artery disease[CAD], stroke, Type 2 diabetes mellitus[T2DM], hypertension[HTN], atrial fibrillation[AF], chronic obstructive pulmonary disease[COPD], dementia, history of cancer, chronic kidney disease[CKD] and pneumonia), overall health status and indicators of multi-morbidities (number of non-cancer illnesses, number of medications prescribed by GP and number of hospitalizations in the past year), indicators of obesity (body mass index [BMI], waist circumference), socioeconomic status (Townsend Deprivation index) and smoking status. Covariates were picked based on their potential relevance to COVID-19 infection and/or its complications. Please refer to Table S1 for the full list and distribution of covariates.

To define history of relevant diseases, we included information from primary care and hospital inpatient data, as well as ICD-10 diagnoses (code 41270) and self-reported illnesses (code 20002). The strategy of integrating all diagnoses were based on instructions from UKBB (https://biobank.ndph.ox.ac.uk/showcase/showcase/docs/first_occurrences_outcomes.pdf).

### Missing covariate data

Missing values of features were imputed with “missRanger”. The program is based on missForest, an iterative imputation approach using random forest. The program has been widely used and has been shown to produce low imputation errors and good performance in predictive models ^17^. The missing rate of each covariate and the OOB (out-of-bag error) of imputation are listed in Table S2.

### COVID-19 infection status

COVID-19 testing data were downloaded from the UKBB data portal (please refer to https://biobank.ndph.ox.ac.uk/showcase/exinfo.cgi?src=COVID19). Briefly, the latest COVID test results were downloaded on 21 Dec 2021 (last update on 30 Oct 2021). COVID-19 test results after the study end-date (30 Sep 2021 for hospital admission and 18 Oct 2021 for mortality) were ignored. COVID-19 infection status was defined by any positive test results, or a diagnosis of COVID-19 (ICD-10 code: U071) from inpatient/mortality records or the code “Y2a3b” within the TPP GP records ((https://biobank.ndph.ox.ac.uk/showcase/coding.cgi?id=8708&nl=1) in the follow-up period.

“Severe” COVID-19 was defined by any case who is hospitalized or whose mortality is due to U07.1. Other non-hospitalized and non-fatal cases are considered as ‘mild’. For hospitalized cases, we required both test result and test origin to be 1 (positive test and inpatient origin). For a small number of cases with initial outpatient origin and positive test result, but changed to inpatient origin within 2 weeks, we still considered these subjects as inpatient cases (i.e., we assume the hospitalization was related to COVID-19).

### Four models of analysis

Four sets of analysis were performed for hospitalization and mortality as outcome separately (Table 1). For model A, it was restricted to test-positive subjects, while for other models, they were based on the whole population. For Model A, we compared severe vs mild COVID-19 cases for post-infection hospital admission and mortalities. For model B, we compared severe COVID-19 cases against those with no known history of infection. For model C, all infected cases were compared against those with no known infection. Finally, ‘mild COVID-19’ were compared against subjects with no known infection in model D.

**Table 1.**
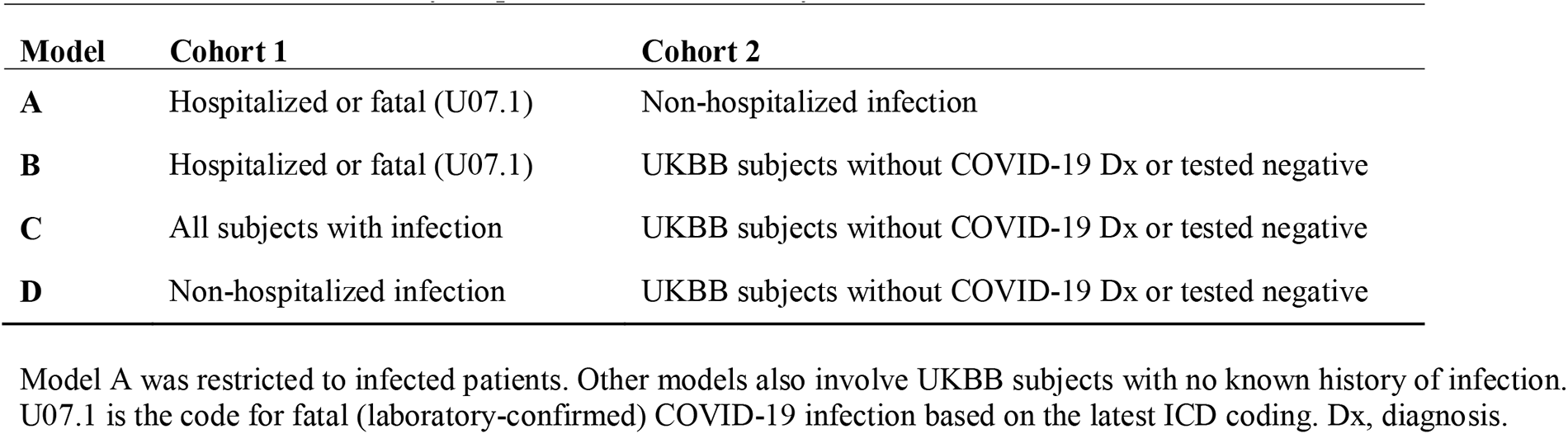
The four sets of analyses performed in this study

### Statistical analysis

Cox and Poisson regression were employed for analysis. Cox regression models time to development of event, while Poisson regression models the incidence rate (with consideration of follow-up time). Briefly, the time to hospitalization or mortality from the studied diseases was treated as outcome in Cox regression, controlling for other covariates. For Poisson regression, presence/absence of the event of interest was considered as outcome, with ‘offset’ specified (=number of days of follow-up) to account for the differences in duration of follow-up for each subject.

For subjects with COVID-19 infection, the first positive date was treated as the start-date (time0) of this cohort study. For subjects free of COVID-19 infection, 31-Jan-2020 were assigned as time0. The end of follow-up was set as the time a participant developed an event of interest or 30 Sep 2021 (for analysis of hospitalization) or 18 Oct 2021 (for analysis of mortality). All statistical analysis were performed by R (version number 3.6.1). The false discovery rate (FDR) approach by Benjamini and Hochberg^18^ was performed to control for multiple testing, which controls the expected proportion of false-positives among the rejected hypotheses.

To avoid convergence issues when large number of predictors are included, we followed the methodology described by Zhao et al.^19^ to further select covariates for inclusion. We first included several ‘default’ variables, including age, sex and general health status (number of hospitalization and medications received in the past year) into the regression model. For the rest of the covariates, univariate testing was performed with each predictor; those with nominally significant associations (p<0.05) with the outcome were selected into the final model. This approach^19^ was shown by theory and simulations to control the proportion of false positive predictors.

### Prior event rate ratio (PERR) adjustment

Although many confounders have been adjusted for, the chance of residual confounding remains. The PERR approach exploits a before-and-after design to minimize the effects of unmeasured confounders^20-22^. The basic idea is to compare the differences in hazard ratios between the exposed and unexposed groups before and after the COVID-19 pandemic, as differences in hospitalization/mortality risks prior to the pandemic can be due to baseline differences (unmeasured confounders) between the two groups.

We may assume that *before* the onset of the pandemic in UK (31 Jan 2020), the hazard ratio of event (e.g. hospitalization due to a specific disease) between the exposed and unexposed groups is HR_before_. Since neither group would have COVID-19 before the baseline, HR_before_ is assumed to reflect baseline differences owing to unmeasured confounders, independent of the effect of COVID-19. Similarly, we define the differences between groups *after* the pandemic onset to be HR_after_. The PERR-adjusted HR, calculated as HR_after_/HR_before_, estimates the effect of COVID-19 accounting for both unmeasured and measured confounders. Equivalently we may compute the difference in Cox regression coefficients. For details please refer to Supplementary Methods.

### Prescription-time distribution matching (PTDM)

In this cohort study, the start-date (time0) for the exposed group is the first date of being tested positive, while an artificial time0 was assigned to the unexposed group. Imbalance of ‘prescription time’ (here ‘prescription’ refers to being infected by COVID-19) distribution may lead to ‘survival bias’. Survival bias may be generated when individuals who die shortly after the start of follow-up might not have a probability of being infected and would only be classified as ‘unexposed’.

In our primary analysis, we excluded the ‘immortal time’ period which gives good estimates of the true coefficients when the exposed group is far smaller than the unexposed group^23^. In addition, if bias remains, the observed effect size (HR) is always smaller than the true HR. Since COVID-19 tends to increase risk of other diseases (HR>1), a smaller HR means that the bias is conservative^23^. Secondly, we also employed the PTDM approach which has been shown to effectively reduce survival bias^24^. In brief, for each person without COVID-19 infection, instead of using the same start-date (31-Jan-2020), an alternative time0 was generated by randomly choosing from the sets of time0 from the COVID-19-exposed group. This ensures a similar distribution of the time0 across both groups (see also Supplementary Methods).

### Other analysis to check for robustness of findings

We performed a variety of different analyses to verify the robustness of our findings under different modelling strategies and assumptions:

1. While we primarily focused on all hospitalizations, we also performed stratified analysis for new-onset and recurrent diseases, as mentioned earlier;
2. for counting the follow-up end date, we primarily consider the last date of available hospitalization/mortality records as the end of follow-up. However, since vaccination may change the risk of sequelae, we also performed analysis limited to the period for which no vaccines were yet available, i.e. setting 8 Dec 2020 (the date of first vaccination in UK) as our study end-date;
3. We performed extra analysis to test associations with subsequent hospitalization and mortality 15 or 30 days *after* being tested positive. The reason is that it is possible that some subjects may be admitted for other disorders and were also tested positive on screening. (However, we note that even in such cases, these patients may be infected by COVID-19 first, and the infection may trigger the disease that leads to the primary admission; this approach may miss such cases).

### Association with hospital admission/mortality stratified by vaccination status and follow-up period

We also conducted further (exploratory) analysis stratified by vaccination status. Briefly, we compared hospitalization rates for subjects who have received at least one dose of vaccine before or after the infection, those who are infected but not vaccinated and those with no known COVID-19. One of our main questions is whether prior vaccination (and to a lesser extent, vaccination after infection) may ameliorate the risk of sequelae, when compared to infected subjects who are not vaccinated.

In another additional analysis, we first tested proportional hazards (PH) assumption for associations that were (nominally) significant (p<0.05). For these outcomes we also examined the effect of COVID-19 on hospitalization/mortality risks as stratified by the follow-up period. Three periods were specified, namely <3 months, 3-6 months and >6 months. The PH assumption were tested again after stratification by follow-up time. Moreover, we also evaluated continuous time-dependent coefficients (https://cran.r-project.org/web/packages/survival/vignettes/timedep.pdf), constructed using the time-transformation functionality of the R function coxph. Briefly, we assume the Cox regression coefficient is time-varying and a linear function of log(*t*), i.e. β(*t*) = *a* + *b* log(*t*). From this we may also calculate the time taken for the regression coefficient (i.e. the adverse effects of infection) to wane to zero. The CI for the “time to no effect” was calculated by the delta method. We hypothesized that the risk would be most prominent in the initial period after infection and would wane afterwards.

## Results

### Overview

In this study, we found that severe COVID-19 was associated with elevated hazards ratios (HR) of hospital admission and mortality due to various diseases involving many body systems. These patterns were consistent in a series of sensitivity analyses, and after PTDM and PERR adjustment.

The four sets of analysis are summarized in Table 1 and the number of subjects for each model is presented in Table 2.

**Table 2.** Number of available subjects in models A to D

| **Model** | **Cohort 1** | **Cohort 2** | **Total** |
| --- | --- | --- | --- |
| **A** | 6484 | 18976 | 25460 |
| **B** | 6484 | 386636 | 393120 |
| **C** | 25460 | 386636 | 412096 |
| **D** | 18976 | 386636 | 405612 |

### Compared to subjects with no known history of COVID-19, association of severe COVID-19 with hospitalization from other disorders (severe disease vs uninfected)

Severe infection was associated with higher hazards of hospitalization and mortality due to a broad array of diseases (median follow-up=608 days). The main results are listed in Table 3 and Fig.2 and full results in Table S3. Here we primarily present the results based on Cox regression model, without further adjustments by PTDM or PERR. Considering all hospitalizations (regardless of history), severe COVID-19 was significantly associated with elevated hazards of hospitalization involving multiple body systems, as described below.

**Fig.2.**
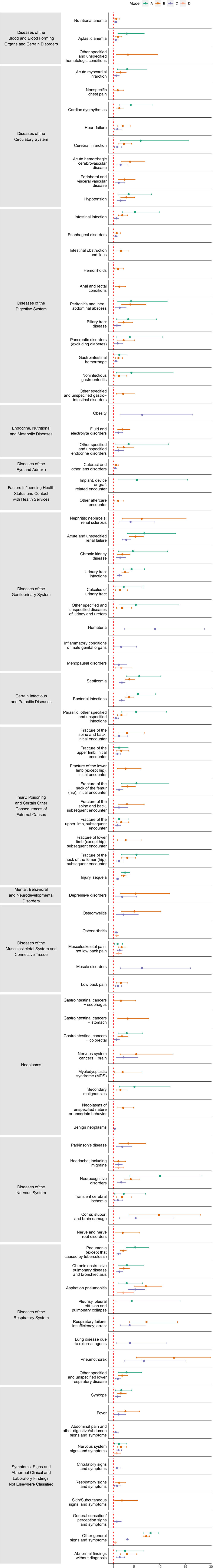
Significant association of COVID-19 with hospitalization due to various diseases, models A to D (containing analysis on all hospitalizations, and without PTDM). The red dashed line stands for the line of no effect (hazard ratio=1). X-axis indicates the HR (hazard ratio) of hospitalization post COVID-19. Y-axis indicates each specific diagnoses category. Confidence intervals are also shown in the figure. We only present the results if the number of events>=5 for both exposed and unexposed groups.

**Table 3.** Compared with subjects with no known history of COVID, the associations of severe COVID-19 with hospitalization due to various disorders, without PTDM adjustment (i.e. model B)

| Outcome | coef | HR | se(coef) | z | P-value | p.adj | Lower CI | Upper CI | CCSR Category Description |
| --- | --- | --- | --- | --- | --- | --- | --- | --- | --- |
| BLD001 | 0.450 | 1.568 | 0.158 | 2.848 | 4.40E-03 | **1.81E-02** | 1.151 | 2.137 | Nutritional anemia |
| BLD003 | 0.635 | 1.887 | 0.166 | 3.836 | 1.25E-04 | **8.82E-04** | 1.364 | 2.611 | Aplastic anemia |
| BLD010 | 1.343 | 3.832 | 0.472 | 2.848 | 4.39E-03 | **1.81E-02** | 1.520 | 9.659 | Other specified and unspecified hematologic conditions |
| CIR009 | 0.862 | 2.369 | 0.198 | 4.347 | 1.38E-05 | **1.34E-04** | 1.606 | 3.494 | Acute myocardial infarction |
| CIR012 | 0.612 | 1.843 | 0.248 | 2.464 | 1.37E-02 | **4.68E-02** | 1.133 | 2.998 | Nonspecific chest pain |
| CIR017 | 0.771 | 2.162 | 0.178 | 4.324 | 1.53E-05 | **1.45E-04** | 1.524 | 3.066 | Cardiac dysrhythmias |
| CIR019 | 1.029 | 2.797 | 0.216 | 4.755 | 1.98E-06 | **2.30E-05** | 1.831 | 4.273 | Heart failure |
| CIR020 | 1.108 | 3.028 | 0.203 | 5.456 | 4.87E-08 | **7.99E-07** | 2.034 | 4.508 | Cerebral infarction |
| CIR021 | 1.446 | 4.244 | 0.265 | 5.450 | 5.04E-08 | **8.17E-07** | 2.524 | 7.138 | Acute hemorrhagic cerebrovascular disease |
| CIR026 | 1.152 | 3.166 | 0.257 | 4.482 | 7.39E-06 | **7.52E-05** | 1.913 | 5.240 | Peripheral and visceral vascular disease |
| CIR031 | 1.256 | 3.512 | 0.191 | 6.570 | 5.02E-11 | **1.16E-09** | 2.415 | 5.109 | Hypotension |
| DIG001 | 1.001 | 2.721 | 0.160 | 6.272 | 3.57E-10 | **7.95E-09** | 1.990 | 3.720 | Intestinal infection |
| DIG004 | 0.486 | 1.626 | 0.200 | 2.436 | 1.49E-02 | **4.98E-02** | 1.100 | 2.405 | Esophageal disorders |
| DIG012 | 0.890 | 2.435 | 0.253 | 3.522 | 4.28E-04 | **2.52E-03** | 1.484 | 3.996 | Intestinal obstruction and ileus |
| DIG014 | 0.641 | 1.899 | 0.222 | 2.889 | 3.86E-03 | **1.63E-02** | 1.229 | 2.935 | Hemorrhoids |
| DIG015 | 0.761 | 2.140 | 0.228 | 3.329 | 8.70E-04 | **4.55E-03** | 1.367 | 3.349 | Anal and rectal conditions |
| DIG016 | 1.450 | 4.264 | 0.271 | 5.354 | 8.60E-08 | **1.30E-06** | 2.508 | 7.251 | Peritonitis and intra-abdominal abscess |
| DIG017 | 1.091 | 2.977 | 0.238 | 4.581 | 4.63E-06 | **5.02E-05** | 1.867 | 4.749 | Biliary tract disease |
| DIG020 | 1.092 | 2.980 | 0.277 | 3.938 | 8.22E-05 | **6.20E-04** | 1.731 | 5.132 | Pancreatic disorders (excluding diabetes) |
| DIG021 | 0.683 | 1.979 | 0.172 | 3.981 | 6.86E-05 | **5.40E-04** | 1.414 | 2.770 | Gastrointestinal hemorrhage |
| DIG022 | 0.719 | 2.052 | 0.284 | 2.530 | 1.14E-02 | **3.99E-02** | 1.176 | 3.581 | Noninfectious gastroenteritis |
| DIG025 | 1.063 | 2.895 | 0.299 | 3.558 | 3.74E-04 | **2.24E-03** | 1.612 | 5.199 | Other specified and unspecified gastrointestinal disorders |
| END011 | 1.010 | 2.746 | 0.211 | 4.793 | 1.65E-06 | **1.96E-05** | 1.817 | 4.150 | Fluid and electrolyte disorders |
| END015 | 1.096 | 2.991 | 0.261 | 4.193 | 2.75E-05 | **2.44E-04** | 1.792 | 4.991 | Other specified and unspecified endocrine disorders |
| EYE002 | 0.402 | 1.495 | 0.155 | 2.602 | 9.26E-03 | **3.37E-02** | 1.104 | 2.024 | Cataract and other lens disorders |
| FAC010 | 0.661 | 1.937 | 0.223 | 2.965 | 3.03E-03 | **1.32E-02** | 1.251 | 2.997 | Other aftercare encounter |
| GEN001 | 1.873 | 6.508 | 0.443 | 4.228 | 2.36E-05 | **2.14E-04** | 2.731 | 15.507 | Nephritis; nephrosis; renal sclerosis |
| GEN002 | 1.665 | 5.285 | 0.131 | 12.713 | 5.02E-37 | **5.07E-35** | 4.089 | 6.832 | Acute and unspecified renal failure |
| GEN003 | 0.997 | 2.711 | 0.230 | 4.336 | 1.45E-05 | **1.40E-04** | 1.727 | 4.256 | Chronic kidney disease |
| GEN004 | 1.182 | 3.262 | 0.109 | 10.862 | 1.76E-27 | **1.44E-25** | 2.635 | 4.037 | Urinary tract infections |
| GEN005 | 0.826 | 2.284 | 0.248 | 3.324 | 8.88E-04 | **4.63E-03** | 1.403 | 3.716 | Calculus of urinary tract |
| GEN006 | 0.991 | 2.693 | 0.266 | 3.718 | 2.01E-04 | **1.34E-03** | 1.597 | 4.540 | Other specified and unspecified diseases of kidney and ureters |
| INF002 | 1.408 | 4.089 | 0.119 | 11.825 | 2.89E-32 | **2.71E-30** | 3.238 | 5.164 | Septicemia |
| INF003 | 1.396 | 4.040 | 0.107 | 13.043 | 7.00E-39 | **7.66E-37** | 3.275 | 4.983 | Bacterial infections |
| INF009 | 0.940 | 2.559 | 0.182 | 5.176 | 2.27E-07 | **3.20E-06** | 1.793 | 3.653 | Parasitic, other specified and unspecified infections |
| INJ002 | 1.293 | 3.643 | 0.334 | 3.871 | 1.08E-04 | **7.80E-04** | 1.893 | 7.010 | Fracture of the spine and back, initial encounter |
| INJ004 | 0.927 | 2.526 | 0.223 | 4.152 | 3.30E-05 | **2.85E-04** | 1.631 | 3.912 | Fracture of the upper limb, initial encounter |
| INJ005 | 1.214 | 3.368 | 0.329 | 3.695 | 2.20E-04 | **1.44E-03** | 1.769 | 6.413 | Fracture of the lower limb (except hip), initial encounter |
| INJ006 | 1.311 | 3.708 | 0.180 | 7.264 | 3.75E-13 | **1.12E-11** | 2.604 | 5.282 | Fracture of the neck of the femur (hip), initial encounter |
| INJ039 | 1.293 | 3.643 | 0.334 | 3.871 | 1.08E-04 | **7.80E-04** | 1.893 | 7.010 | Fracture of the spine and back, subsequent encounter |
| INJ041 | 0.927 | 2.526 | 0.223 | 4.152 | 3.30E-05 | **2.85E-04** | 1.631 | 3.912 | Fracture of the upper limb, subsequent encounter |
| INJ042 | 1.214 | 3.368 | 0.329 | 3.695 | 2.20E-04 | **1.44E-03** | 1.769 | 6.413 | Fracture of lower limb (except hip), subsequent encounter |
| INJ043 | 1.311 | 3.708 | 0.180 | 7.264 | 3.75E-13 | **1.12E-11** | 2.604 | 5.282 | Fracture of the neck of the femur (hip), subsequent encounter |
| INJ073 | 1.094 | 2.985 | 0.076 | 14.342 | 1.19E-46 | **1.57E-44** | 2.571 | 3.466 | Injury, sequela |
| MBD002 | 1.673 | 5.329 | 0.411 | 4.069 | 4.73E-05 | **3.93E-04** | 2.380 | 11.929 | Depressive disorders |
| MUS002 | 1.628 | 5.095 | 0.358 | 4.544 | 5.51E-06 | **5.79E-05** | 2.524 | 10.283 | Osteomyelitis |
| MUS010 | 0.953 | 2.593 | 0.144 | 6.617 | 3.68E-11 | **8.77E-10** | 1.955 | 3.439 | Musculoskeletal pain, not low back pain |
| MUS038 | 0.894 | 2.444 | 0.212 | 4.223 | 2.41E-05 | **2.15E-04** | 1.615 | 3.701 | Low back pain |
| NEO013 | 1.331 | 3.787 | 0.374 | 3.562 | 3.69E-04 | **2.22E-03** | 1.820 | 7.879 | Gastrointestinal cancers - stomach |
| NEO015 | 0.980 | 2.664 | 0.188 | 5.204 | 1.95E-07 | **2.78E-06** | 1.842 | 3.853 | Gastrointestinal cancers - colorectal |
| NEO048 | 1.694 | 5.443 | 0.430 | 3.943 | 8.05E-05 | **6.11E-04** | 2.345 | 12.636 | Nervous system cancers - brain |
| NEO070 | 0.863 | 2.370 | 0.220 | 3.925 | 8.66E-05 | **6.42E-04** | 1.540 | 3.646 | Secondary malignancies |
| NEO072 | 1.069 | 2.912 | 0.266 | 4.012 | 6.03E-05 | **4.83E-04** | 1.727 | 4.908 | Neoplasms of unspecified nature or uncertain behavior |
| NVS004 | 1.355 | 3.877 | 0.324 | 4.187 | 2.83E-05 | **2.49E-04** | 2.056 | 7.312 | Parkinson`s disease |
| NVS010 | 0.677 | 1.968 | 0.274 | 2.472 | 1.34E-02 | **4.60E-02** | 1.150 | 3.366 | Headache; including migraine |
| NVS011 | 1.473 | 4.364 | 0.175 | 8.429 | 3.50E-17 | **1.39E-15** | 3.098 | 6.146 | Neurocognitive disorders |
| NVS012 | 0.974 | 2.649 | 0.269 | 3.629 | 2.85E-04 | **1.79E-03** | 1.565 | 4.484 | Transient cerebral ischemia |
| NVS013 | 2.287 | 9.848 | 0.454 | 5.039 | 4.68E-07 | **6.03E-06** | 4.045 | 23.972 | Coma; stupor; and brain damage |
| NVS017 | 1.031 | 2.804 | 0.392 | 2.630 | 8.53E-03 | **3.14E-02** | 1.301 | 6.046 | Nerve and nerve root disorders |
| RSP002 | 1.060 | 2.886 | 0.098 | 10.799 | 3.49E-27 | **2.70E-25** | 2.381 | 3.498 | Pneumonia (except that caused by tuberculosis) |
| RSP008 | 1.103 | 3.013 | 0.159 | 6.939 | 3.96E-12 | **1.06E-10** | 2.206 | 4.114 | Chronic obstructive pulmonary disease and bronchiectasis |
| RSP010 | 1.992 | 7.332 | 0.181 | 11.008 | 3.50E-28 | **3.06E-26** | 5.142 | 10.453 | Aspiration pneumonitis |
| RSP012 | 2.007 | 7.442 | 0.303 | 6.623 | 3.53E-11 | **8.58E-10** | 4.109 | 13.480 | Respiratory failure; insufficiency; arrest |
| RSP014 | 2.549 | 12.799 | 0.432 | 5.896 | 3.71E-09 | **7.07E-08** | 5.485 | 29.867 | Pneumothorax |
| RSP016 | 0.991 | 2.694 | 0.167 | 5.926 | 3.11E-09 | **6.07E-08** | 1.941 | 3.739 | Other specified and unspecified lower respiratory disease |
| SYM001 | 0.898 | 2.455 | 0.181 | 4.965 | 6.86E-07 | **8.58E-06** | 1.722 | 3.500 | Syncope |
| SYM002 | 1.207 | 3.343 | 0.305 | 3.963 | 7.40E-05 | **5.70E-04** | 1.840 | 6.073 | Fever |
| SYM010 | 0.919 | 2.508 | 0.181 | 5.075 | 3.87E-07 | **5.09E-06** | 1.758 | 3.577 | Nervous system signs and symptoms |
| SYM013 | 0.745 | 2.107 | 0.244 | 3.051 | 2.28E-03 | **1.05E-02** | 1.305 | 3.401 | Respiratory signs and symptoms |
| SYM014 | 0.978 | 2.660 | 0.393 | 2.492 | 1.27E-02 | **4.39E-02** | 1.232 | 5.741 | Skin/Subcutaneous signs and symptoms |
| SYM016 | 1.999 | 7.383 | 0.042 | 47.386 | 0.00E+00 | **0.00E+00** | 6.797 | 8.019 | Other general signs and symptoms |
| SYM017 | 1.282 | 3.605 | 0.218 | 5.883 | 4.03E-09 | **7.56E-08** | 2.352 | 5.526 | Abnormal findings without diagnosis |

Coef, regression coefficient from Cox model. HR, hazard ratio; se, standard error; p.adj, FDR-adjusted p-value; Lower and upper CI represent the lower and upper 95% confidence interval of HR.

#### Respiratory diseases

Elevated hazards of hospitalization due to respiratory conditions after severe COVID-19 infection were observed. Increased risk of hospital admission was observed for pneumonia (except that caused by tuberculosis) (Hazard ratio [HR]=2.886 (95% Confidence Interval: 2.381-3.498); FDR-adjusted p-value (p.adj)=2.70e-25) (figures in the parentheses organized in the same way below), chronic obstructive pulmonary disease and bronchiectasis (3.013 (2.206-4.114); p.adj=1.06e-10), aspiration pneumonitis (7.332 (5.142-10.453); p.adj=3.06e-26), respiratory failure/insufficiency/arrest (7.442 (4.109-13.480); p.adj=8.58e-10), pneumothorax (12.799 (5.485-29.867); p.adj=7.07e-08), and other specified and unspecified lower respiratory disease (2.694 (1.941-3.739); p.adj=6.07e-08), which includes interstitial pulmonary disorder, hypostatic pneumonia without specified organism, rheumatoid lung disease, etc.

#### Cardiovascular diseases

We observed increased hazards of hospital admission from several cardiovascular/cerebrovascular conditions, including acute myocardial infarction (AMI) (2.369 (1.606-3.494); p.adj=1.34e-04), nonspecific chest pain (1.843 (1.133-2.998); p.adj=4.68e-02), cardiac dysrhythmias (2.162 (1.524-3.066); p.adj=1.45e-04), heart failure (2.797 (1.831-4.273); p.adj=2.30e-05), cerebral infarction (3.028 (2.034-4.508); p.adj=7.99e-07), acute hemorrhagic cerebrovascular disease (4.244 (2.524-7.138); p.adj=8.17e-07), peripheral and visceral vascular disease (3.166 (1.913-5.240); p.adj=7.52e-05) and hypotension (3.512 (2.415-5.109); p.adj=1.16e-09).

#### Genitourinary diseases

There was evidence for increased hazards of hospitalization from genitourinary disorders, including the following conditions: nephritis/nephrosis/renal sclerosis (6.508 (2.731-15.507); p.adj=2.14e-04), acute and unspecified renal failure (5.285 (4.089-6.832); p.adj=5.07e-35), chronic kidney disease (CKD) (2.711 (1.727-4.256); p.adj=1.40e-04), urinary tract infection (UTI) (3.262 (2.635-4.037); p.adj=1.44e-25), calculus of urinary tract (2.284 (1.403-3.716); p.adj=4.63e-03), other specified and unspecified diseases of kidney and ureters (e.g. hydronephrosis, obstructive and reflux uropathy, cyst) (2.693 (1.597-4.540); p.adj=1.34e-03).

#### Nervous system disorders

Increased hazards of hospitalization from neurological disorders were evident and included Parkinson’s disease (3.877 (2.056-7.312); p.adj=2.49e-04), headache (including migraine) (1.968 (1.150-3.366); p.adj=4.60e-02), neurocognitive disorders (4.364 (3.098-6.146); p.adj=1.39e-15), transient cerebral ischemia (2.649 (1.565-4.484); p.adj=1.79e-03), coma, stupor and brain damage (9.848 (4.045-23.972); p.adj=6.03e-06) and nerve and nerve root disorders (2.804 (1.301-6.046); p.adj=3.14e-02).

#### Digestive conditions

Our results showed increased hazards of hospitalization due to digestive disorders after severe COVID-19, including oesophageal disorders (1.626 (1.100-2.405); p.adj=4.98e-02), pancreatic disorders (excluding diabetes) (2.980 (1.731-5.132); p.adj=6.20e-04), hemorrhage of digestive tract (1.979 (1.414-2.770); p.adj=5.40e-04), infectious and non-infectious diseases of digestive tract (2.052 (1.176-3.581); p.adj=3.99e-02). Elevated hazards of hospitalization due to other specified and unspecified gastrointestinal disorders (2.895 (1.612-5.199); p.adj=2.24e-03) was also observed.

#### Musculoskeletal disorders

Severe COVID-19 were associated with increasedhazards of hospitalization from osteomyelitis (5.095 (2.524-10.283); p.adj=5.79e-05), musculoskeletal pain (2.593 (1.955-3.439); p.adj=8.77e-10) and low back pain (2.444 (1.615-3.701); p.adj=2.15e-04). Higher hospitalization risks from initial encounters of fracture were observed, including fracture of the spine and back (3.643 (1.893-7.010); p.adj=7.80e-04), the upper limb (2.526 (1.631-3.912); p.adj=2.85e-04), the lower limb (except hip (3.368 (1.769-6.413); p.adj=1.44e-03) and the neck of the femur (hip), (3.708 (2.604-5.282); p.adj=1.12e-11). High hazards of hospital admission were also seen in subsequent encounter of above-mentioned fracture and other injury sequelae, like dislocation, sprain, etc.

#### Neoplasms

History of severe COVID-19 was associated with increased hazards of hospitalization from several neoplasms, including stomach cancers (3.787 (1.820-7.879); p.adj=2.22e-03), colorectal cancers (2.664 (1.842-3.853); p.adj=2.78e-06), brain cancers (5.443 (2.345-12.636); p.adj=6.11e-04), secondary malignancies (2.370 (1.540-3.646); p.adj=6.42e-04) and neoplasms of unspecified nature or uncertain behaviour (2.912 (1.727-4.908); p.adj=4.83e-04).

#### Other infections

History of severe COVID-19 was associated with increased hazards of hospitalization from septicemia (4.089 (3.238-5.164); p.adj=2.71e-30), bacterial infections (4.040 (3.275-4.983); p.adj=7.66e-37), as well as parasitic and other unspecified infections (2.559 (1.793-3.653); p.adj=3.20e-06).

#### Other sequelae

We also observed increased hazards of hospitalization from nutritional anemia (1.568 (1.151-2.137); p.adj=1.81e-02), aplastic anemia (1.887 (1.364-2.611); p.adj=8.82e-04), metabolic disorders, like fluid and electrolyte disorders (2.746 (1.817-4.150); p.adj=1.96e-05), depressive disorders (5.329 (2.380-11.929); p.adj=3.93e-04), and general poor wellbeing, such as syncope (2.455 (1.722-3.500); p.adj=8.58e-06), fever (3.343 (1.840-6.073); p.adj=5.70e-04), and other general signs and symptoms (7.383 (6.797-8.019); p.adj<9.60e-231), abnormal findings without diagnosis (3.605(2.352-5.526); p.adj=7.56e-08).

When we restricted the outcome to hospitalization from *new-onset or recurrent* diseases, most of our results were similar (see Fig.S1). Higher risks of hospitalization from a few additional disorders were found when we performed analysis on hospitalizations from new-onset diseases, including coronary atherosclerosis and other heart diseases (2.248 (1.195-4.229); p.adj=4.19e-02), benign neoplasms (1.576 (1.121-2.215); p.adj=3.24e-02), and other specified hereditary and degenerative nervous system conditions (4.763 (1.851-12.257); p.adj=6.10e-03).

We also repeated the analysis using Poisson regression which models the incidence rate of hospitalization. The results were similar to those from Cox regression, with similar significant associations observed (Table S3).

### Compared to subjects with mild COVID-19, association of severe COVID-19 with hospitalization from other disorders (severe vs mild infection)

Here we compared the risk of hospitalization (from other disorders) of severe vs mild cases *within the infected* individuals (Table 4 and Fig.2). Compared with mild COVID-19, higher hazards of hospitalization from multiple diseases were observed after severe infection (median follow-up: 248 days), including for example, hospitalization from aplastic anaemia (hazard ratio [HR]= 3.606 (95% CI: 1.854-7.014); FDR-adjusted p-value (p.adj)= 1.08e-03), acute myocardial infarction [AMI] (HR=3.674 (1.788-7.549); p.adj=2.36e-03), cardiac dysrhythmias (HR=4.364 (2.232-8.531); p.adj=1.54e-04), cerebral infarction (HR=6.307 (2.331-17.061); p.adj= 1.79e-03), hypotension (HR=3.965 (1.867-8.423); p.adj= 2.07e-03), acute and unspecified renal failure (HR=7.009 (3.744-13.119); p.adj= 2.38e-08), chronic kidney disease [CKD] (HR=4.769 (1.970-11.542); p.adj= 3.01e-03), urinary tract infections [UTI] (HR=4.533 (2.915-7.048); p.adj= 4.85e-10), calculus of urinary tract (HR=2.992 (1.321-6.777); p.adj= 3.16e-02), neurocognitive disorders (HR=10.081 (4.201-24.193); p.adj= 3.21e-06), pneumonia (except that caused by tuberculosis) (HR=5.226 (3.446-7.924); p.adj= 2.46e-13), chronic obstructive pulmonary disease and bronchiectasis [COPD] (HR=3.633 (1.904-6.935); p.adj= 6.71e-04), and aspiration pneumonitis (HR=3.573 (1.921-6.644); p.adj= 4.66e-04). We also observed increased risk of hospitalization from several digestive disorders, infectious conditions, injury and general signs/symptoms, which are listed in Table 4.

**Table 4.** Compared to subjects with mild (non-hospitalized) COVID, the associations of severe COVID-19 with hospitalization due to various disorders, without PTDM adjustment (i.e. model A)

| Outcome | coef | HR | se(coef) | z | P-value | p.adj | Lower CI | Upper CI | CCSR Category Description |
| --- | --- | --- | --- | --- | --- | --- | --- | --- | --- |
| BLD003 | 1.283 | 3.606 | 0.339 | 3.779 | 1.57E-04 | **1.08E-03** | 1.854 | 7.014 | Aplastic anemia |
| CIR009 | 1.301 | 3.674 | 0.367 | 3.542 | 3.98E-04 | **2.36E-03** | 1.788 | 7.549 | Acute myocardial infarction |
| CIR017 | 1.473 | 4.364 | 0.342 | 4.307 | 1.65E-05 | **1.54E-04** | 2.232 | 8.531 | Cardiac dysrhythmias |
| CIR020 | 1.842 | 6.307 | 0.508 | 3.627 | 2.87E-04 | **1.79E-03** | 2.331 | 17.061 | Cerebral infarction |
| CIR031 | 1.378 | 3.965 | 0.384 | 3.584 | 3.39E-04 | **2.07E-03** | 1.867 | 8.423 | Hypotension |
| DIG001 | 1.656 | 5.237 | 0.330 | 5.011 | 5.42E-07 | **6.91E-06** | 2.740 | 10.008 | Intestinal infection |
| DIG016 | 1.489 | 4.432 | 0.486 | 3.065 | 2.17E-03 | **1.01E-02** | 1.711 | 11.483 | Peritonitis and intra-abdominal abscess |
| DIG017 | 1.361 | 3.899 | 0.449 | 3.031 | 2.44E-03 | **1.10E-02** | 1.617 | 9.400 | Biliary tract disease |
| DIG020 | 1.428 | 4.172 | 0.471 | 3.030 | 2.44E-03 | **1.10E-02** | 1.656 | 10.508 | Pancreatic disorders (excluding diabetes) |
| DIG021 | 0.768 | 2.156 | 0.271 | 2.838 | 4.54E-03 | **1.86E-02** | 1.268 | 3.665 | Gastrointestinal hemorrhage |
| DIG022 | 1.504 | 4.500 | 0.527 | 2.854 | 4.32E-03 | **1.81E-02** | 1.602 | 12.643 | Noninfectious gastroenteritis |
| END015 | 1.375 | 3.955 | 0.556 | 2.474 | 1.33E-02 | **4.58E-02** | 1.331 | 11.756 | Other specified and unspecified endocrine disorders |
| FAC009 | 1.719 | 5.582 | 0.548 | 3.137 | 1.71E-03 | **8.30E-03** | 1.906 | 16.342 | Implant, device or graft related encounter |
| GEN002 | 1.947 | 7.009 | 0.320 | 6.088 | 1.14E-09 | **2.38E-08** | 3.744 | 13.119 | Acute and unspecified renal failure |
| GEN003 | 1.562 | 4.769 | 0.451 | 3.464 | 5.32E-04 | **3.01E-03** | 1.970 | 11.542 | Chronic kidney disease |
| GEN004 | 1.511 | 4.533 | 0.225 | 6.711 | 1.94E-11 | **4.85E-10** | 2.915 | 7.048 | Urinary tract infections |
| GEN005 | 1.096 | 2.992 | 0.417 | 2.627 | 8.62E-03 | **3.16E-02** | 1.321 | 6.777 | Calculus of urinary tract |
| GEN006 | 1.678 | 5.353 | 0.481 | 3.486 | 4.91E-04 | **2.81E-03** | 2.084 | 13.748 | Other specified and unspecified diseases of kidney and ureters |
| INF002 | 1.798 | 6.039 | 0.268 | 6.709 | 1.96E-11 | **4.85E-10** | 3.571 | 10.212 | Septicemia |
| INF003 | 1.758 | 5.801 | 0.237 | 7.434 | 1.06E-13 | **3.46E-12** | 3.649 | 9.222 | Bacterial infections |
| INF009 | 1.693 | 5.438 | 0.374 | 4.533 | 5.81E-06 | **6.05E-05** | 2.615 | 11.310 | Parasitic, other specified and unspecified infections |
| INJ006 | 1.703 | 5.489 | 0.392 | 4.349 | 1.37E-05 | **1.34E-04** | 2.548 | 11.824 | Fracture of the neck of the femur (hip), initial encounter |
| INJ043 | 1.703 | 5.489 | 0.392 | 4.349 | 1.37E-05 | **1.34E-04** | 2.548 | 11.824 | Fracture of the neck of the femur (hip), subsequent encounter |
| INJ073 | 1.184 | 3.269 | 0.131 | 9.020 | 1.87E-19 | **8.49E-18** | 2.527 | 4.228 | Injury, sequela |
| MUS010 | 0.593 | 1.810 | 0.215 | 2.757 | 5.83E-03 | **2.34E-02** | 1.187 | 2.760 | Musculoskeletal pain, not low back pain |
| NEO015 | 1.279 | 3.593 | 0.318 | 4.023 | 5.74E-05 | **4.66E-04** | 1.927 | 6.701 | Gastrointestinal cancers - colorectal |
| NEO070 | 1.628 | 5.095 | 0.439 | 3.706 | 2.11E-04 | **1.40E-03** | 2.153 | 12.056 | Secondary malignancies |
| NVS011 | 2.311 | 10.081 | 0.447 | 5.173 | 2.30E-07 | **3.21E-06** | 4.201 | 24.193 | Neurocognitive disorders |
| RSP002 | 1.654 | 5.226 | 0.212 | 7.786 | 6.93E-15 | **2.46E-13** | 3.446 | 7.924 | Pneumonia (except that caused by tuberculosis) |
| RSP008 | 1.290 | 3.633 | 0.330 | 3.912 | 9.15E-05 | **6.71E-04** | 1.904 | 6.935 | Chronic obstructive pulmonary disease and bronchiectasis |
| RSP010 | 1.273 | 3.573 | 0.317 | 4.023 | 5.74E-05 | **4.66E-04** | 1.921 | 6.644 | Aspiration pneumonitis |
| RSP011 | 1.516 | 4.552 | 0.573 | 2.645 | 8.17E-03 | **3.07E-02** | 1.481 | 13.996 | Pleurisy, pleural effusion and pulmonary collapse |
| RSP016 | 1.270 | 3.559 | 0.307 | 4.137 | 3.52E-05 | **3.00E-04** | 1.950 | 6.495 | Other specified and unspecified lower respiratory disease |
| SYM001 | 0.933 | 2.542 | 0.297 | 3.140 | 1.69E-03 | **8.24E-03** | 1.420 | 4.550 | Syncope |
| SYM010 | 0.729 | 2.073 | 0.279 | 2.614 | 8.95E-03 | **3.27E-02** | 1.200 | 3.582 | Nervous system signs and symptoms |
| SYM016 | 2.108 | 8.231 | 0.090 | 23.440 | 1.65E-121 | **3.62E-119** | 6.901 | 9.817 | Other general signs and symptoms |
| SYM017 | 1.192 | 3.294 | 0.379 | 3.146 | 1.65E-03 | **8.13E-03** | 1.567 | 6.921 | Abnormal findings without diagnosis |

### Compared to subjects with no known history of COVID-19, association of mild COVID-19 with hospitalization from other disorders (mild infection vs uninfected)

We found from this analysis (model D) that history of ‘mild’ (i.e. non-hospitalized) COVID-19 were associated increased hazards of hospital admission due to musculoskeletal pain (excluding low back pain) (HR=1.937 (1.463-2.566); p.adj=4.29e-05), aspiration pneumonitis (HR=2.933 (1.716-5.012); p.adj=6.25e-04), and other general signs and symptoms (like edema, abnormal weight loss, anorexia, and etc.) (HR=1.353 (1.163-1.574); p.adj= 6.59e-04), as shown in Table S3 and Fig.2.

Higher hazards of all-cause mortality (1.237 (1.037-1.476); p.adj=2.27e-02) and mortality due to neurocognitive disorders (9.100 (5.590-14.816); p.adj=7.33e-18) were also observed.

### Compared with subjects with no known history of COVID-19, association of any COVID-19 infection with hospitalization from other disorders (any infection vs uninfected)

The results from this analysis (model C) were largely similar to those from model B. Details can be found in Table S3 and Fig.2. In addition to findings from model B, we also found increased hazards of hospitalization from essential hypertension (3.061 (1.397-6.705); p.adj=2.09e-02), obesity (6.609 (2.263-19.298); p.adj=3.10e-03), hematuria (9.131 (3.184-26.186); p.adj=3.27e-04), osteoarthritis (1.515 (1.267-1.813); p.adj=5.75e-05), muscle disorders (6.564 (2.390-18.022); p.adj=1.66e-03), lung disease due to external agents (4.211 (1.558-11.384); p.adj=1.88e-02), abdominal pain and other digestive/abdomen signs and symptoms (1.591 (1.098-2.306); p.adj=4.79e-02) and circulatory signs and symptoms (1.645 (1.104-2.451); p.adj=4.88e-02).

### Association of severe COVID-19 with hospitalization from other disorders, with PTDM adjustment

Results with PTDM adjustment are listed in Table S3 and Fig.S2. Since PTDM pushed the start-date of the unexposed to a later date, the number of events was smaller for the unexposed. We relaxed our results presenting criteria (cell count>=3 for at least one cell). Under the PTDM adjustment, increased risks of hospitalization were still observed for multiple pulmonary disorders, such as pneumonia (except that caused by tuberculosis) (3.612 (2.944-4.432); p.adj=1.75e-32), COPD (4.325 (3.102-6.029); p.adj=4.13e-16), aspiration pneumonitis (7.796 (5.303-11.460); p.adj=2.00e-23), respiratory failure; insufficiency; arrest (8.559 (4.559-16.070); p.adj=8.64e-10), pneumothorax (19.199 (7.665-48.088); p.adj=9.23e-09), lung disease due to external agents (8.660 (2.210-33.939); p.adj=1.55e-02).

In addition, increased hazards of hospital admission from cardiovascular and several other diseases were observed, such as AMI (2.167 (1.459-3.217); p.adj=1.56e-03), other and ill-defined heart disease (10.610 (2.696-41.752); p.adj=7.15e-03), cardiac dysrhythmias (2.195 (1.539-3.130); p.adj=2.27e-04), heart failure (2.688 (1.731-4.173); p.adj=1.74e-04), cerebral infarction (3.274 (2.167-4.945); p.adj=4.94e-07), acute hemorrhagic cerebrovascular disease (4.251 (2.480-7.288); p.adj=3.36e-06), peripheral and visceral vascular disease (3.986 (2.354-6.747); p.adj=5.93e-06), hypotension (3.002 (2.034-4.431); p.adj=8.06e-07), acute phlebitis; thrombophlebitis and thromboembolism (8.753 (3.037-25.224); p.adj=7.91e-04), nephritis; nephrosis; renal sclerosis (7.471 (2.967-18.814); p.adj=3.07e-04), acute and unspecified renal failure (5.321 (4.051-6.989); p.adj=5.60e-31), chronic kidney disease (5.915 (3.766-9.291); p.adj=6.69e-13); mental disorders (schizophrenia spectrum and other psychotic disorders (9.199 (3.108-27.224); p.adj=8.14e-04)), and rheumatoid arthritis/related disease (2.992 (1.512-5.923); p.adj=1.34e-02). Elevated risks of hospitalization due to some other infectious diseases, injury, neoplasms, and general poor well-being were also observed.

### Association results with PERR adjustment

The primary results with PERR adjustment (with prior exposure period restricted to 30-Jan-2018 to 30-Jan-2020) are listed in Table 5 and Fig.3.

**Fig.3.**
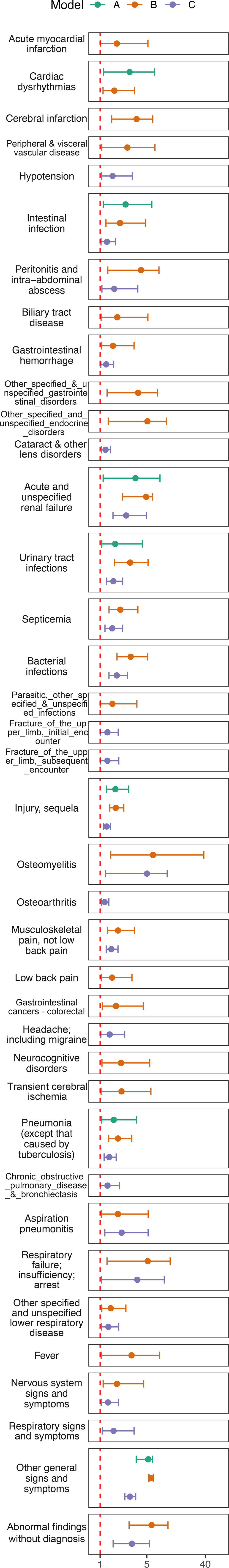
Association of hospitalization with COVID-19 with PERR adjustment. Associations COVID-19 with hospitalization from other diseases with prior event rate ratio (PERR) adjustment. Significant results in the primary analysis were further selected for analysis with PERR adjustment. The red dashed line stands for the line of no effect. We only present the results if the number of events>=5 for both exposed and unexposed groups.

**Table 5.** The associations of any hospitalization with severe COVID-19 due to various disorders, *with PERR adjustment*.

| Outcome | Model | beta_diff | boot_se | zval_final | pval_final | HR | CI_lower | CI_upper | CCSR.Category.Description |
| --- | --- | --- | --- | --- | --- | --- | --- | --- | --- |
| CIR009 | B | 0.889 | 0.431 | 2.063 | 3.91E-02 | 2.432 | 1.045 | 5.658 | Acute myocardial infarction |
| CIR017 | A | 1.260 | 0.514 | 2.452 | 1.42E-02 | 3.524 | 1.288 | 9.646 | Cardiac dysrhythmias |
| CIR017 | B | 0.797 | 0.293 | 2.718 | 6.57E-03 | 2.219 | 1.249 | 3.941 | Cardiac dysrhythmias |
| CIR020 | B | 1.416 | 0.372 | 3.810 | 1.39E-04 | 4.120 | 1.989 | 8.535 | Cerebral infarction |
| CIR026 | B | 1.203 | 0.552 | 2.180 | 2.93E-02 | 3.331 | 1.129 | 9.825 | Peripheral and visceral vascular disease |
| CIR031 | C | 0.727 | 0.303 | 2.396 | 1.66E-02 | 2.069 | 1.142 | 3.751 | Hypotension |
| DIG001 | A | 1.157 | 0.469 | 2.466 | 1.37E-02 | 3.180 | 1.268 | 7.974 | Intestinal infection |
| DIG001 | B | 0.996 | 0.302 | 3.301 | 9.63E-04 | 2.707 | 1.499 | 4.889 | Intestinal infection |
| DIG001 | C | 0.460 | 0.197 | 2.339 | 1.94E-02 | 1.584 | 1.077 | 2.329 | Intestinal infection |
| DIG016 | B | 1.505 | 0.511 | 2.944 | 3.24E-03 | 4.503 | 1.653 | 12.262 | Peritonitis and intra-abdominal abscess |
| DIG016 | C | 0.793 | 0.330 | 2.401 | 1.64E-02 | 2.211 | 1.157 | 4.225 | Peritonitis and intra-abdominal abscess |
| DIG017 | B | 0.903 | 0.421 | 2.148 | 3.17E-02 | 2.468 | 1.082 | 5.627 | Biliary tract disease |
| DIG021 | B | 0.742 | 0.316 | 2.345 | 1.90E-02 | 2.100 | 1.130 | 3.904 | Gastrointestinal hemorrhage |
| DIG021 | C | 0.417 | 0.180 | 2.313 | 2.07E-02 | 1.517 | 1.066 | 2.160 | Gastrointestinal hemorrhage |
| DIG025 | B | 1.448 | 0.500 | 2.896 | 3.78E-03 | 4.255 | 1.597 | 11.336 | Other specified and unspecified gastrointestinal disorders |
| END015 | B | 1.672 | 0.584 | 2.864 | 4.19E-03 | 5.323 | 1.695 | 16.716 | Other specified and unspecified endocrine disorders |
| EYE002 | C | 0.380 | 0.133 | 2.866 | 4.16E-03 | 1.463 | 1.128 | 1.898 | Cataract and other lens disorders |
| GEN002 | A | 1.393 | 0.592 | 2.351 | 1.87E-02 | 4.025 | 1.260 | 12.854 | Acute and unspecified renal failure |
| GEN002 | B | 1.597 | 0.269 | 5.947 | 2.73E-09 | 4.941 | 2.918 | 8.364 | Acute and unspecified renal failure |
| GEN002 | C | 1.172 | 0.218 | 5.367 | 8.00E-08 | 3.229 | 2.105 | 4.955 | Acute and unspecified renal failure |
| GEN004 | A | 0.831 | 0.356 | 2.334 | 1.96E-02 | 2.296 | 1.142 | 4.614 | Urinary tract infections |
| GEN004 | B | 1.272 | 0.240 | 5.294 | 1.20E-07 | 3.569 | 2.228 | 5.716 | Urinary tract infections |
| GEN004 | C | 0.759 | 0.162 | 4.683 | 2.83E-06 | 2.135 | 1.554 | 2.933 | Urinary tract infections |
| INF002 | B | 1.004 | 0.224 | 4.491 | 7.10E-06 | 2.730 | 1.761 | 4.231 | Septicemia |
| INF002 | C | 0.712 | 0.185 | 3.852 | 1.17E-04 | 2.037 | 1.418 | 2.926 | Septicemia |
| INF003 | B | 1.284 | 0.199 | 6.439 | 1.20E-10 | 3.610 | 2.442 | 5.335 | Bacterial infections |
| INF003 | C | 0.887 | 0.164 | 5.396 | 6.80E-08 | 2.427 | 1.759 | 3.350 | Bacterial infections |
| INF009 | B | 0.719 | 0.359 | 2.001 | 4.54E-02 | 2.052 | 1.015 | 4.148 | Parasitic, other specified and unspecified infections |
| INJ004 | C | 0.486 | 0.229 | 2.121 | 3.39E-02 | 1.626 | 1.038 | 2.548 | Fracture of the upper limb, initial encounter |
| INJ041 | C | 0.486 | 0.239 | 2.031 | 4.22E-02 | 1.626 | 1.017 | 2.599 | Fracture of the upper limb, subsequent encounter |
| INJ073 | A | 0.836 | 0.205 | 4.069 | 4.72E-05 | 2.306 | 1.542 | 3.449 | Injury, sequela |
| INJ073 | B | 0.850 | 0.130 | 6.516 | 7.21E-11 | 2.339 | 1.812 | 3.021 | Injury, sequela |
| INJ073 | C | 0.445 | 0.097 | 4.580 | 4.66E-06 | 1.561 | 1.290 | 1.888 | Injury, sequela |
| MUS002 | B | 2.156 | 0.770 | 2.800 | 5.12E-03 | 8.640 | 1.909 | 39.096 | Osteomyelitis |
| MUS002 | C | 1.616 | 0.626 | 2.581 | 9.86E-03 | 5.033 | 1.475 | 17.173 | Osteomyelitis |
| MUS006 | C | 0.328 | 0.118 | 2.776 | 5.50E-03 | 1.388 | 1.101 | 1.750 | Osteoarthritis |
| MUS010 | B | 0.929 | 0.225 | 4.128 | 3.65E-05 | 2.531 | 1.629 | 3.934 | Musculoskeletal pain, not low back pain |
| MUS010 | C | 0.674 | 0.130 | 5.185 | 2.16E-07 | 1.962 | 1.521 | 2.531 | Musculoskeletal pain, not low back pain |
| MUS038 | B | 0.704 | 0.312 | 2.258 | 2.40E-02 | 2.023 | 1.097 | 3.728 | Low back pain |
| NEO015 | B | 0.859 | 0.351 | 2.450 | 1.43E-02 | 2.360 | 1.187 | 4.692 | Gastrointestinal cancers - colorectal |
| NVS010 | C | 0.598 | 0.272 | 2.195 | 2.82E-02 | 1.818 | 1.066 | 3.102 | Headache; including migraine |
| NVS011 | B | 1.025 | 0.447 | 2.293 | 2.19E-02 | 2.786 | 1.160 | 6.689 | Neurocognitive disorders |
| NVS012 | B | 1.040 | 0.491 | 2.117 | 3.42E-02 | 2.829 | 1.080 | 7.409 | Transient cerebral ischemia |
| RSP002 | A | 0.775 | 0.328 | 2.359 | 1.83E-02 | 2.170 | 1.140 | 4.129 | Pneumonia (except that caused by tuberculosis) |
| RSP002 | B | 0.927 | 0.195 | 4.755 | 1.98E-06 | 2.528 | 1.725 | 3.705 | Pneumonia (except that caused by tuberculosis) |
| RSP002 | C | 0.579 | 0.146 | 3.981 | 6.86E-05 | 1.785 | 1.342 | 2.374 | Pneumonia (except that caused by tuberculosis) |
| RSP008 | C | 0.492 | 0.246 | 2.000 | 4.55E-02 | 1.635 | 1.010 | 2.648 | Chronic obstructive pulmonary disease and bronchiectasis |
| RSP010 | B | 0.925 | 0.424 | 2.182 | 2.91E-02 | 2.521 | 1.099 | 5.786 | Aspiration pneumonitis |
| RSP010 | C | 1.045 | 0.360 | 2.905 | 3.68E-03 | 2.842 | 1.405 | 5.752 | Aspiration pneumonitis |
| RSP012 | B | 1.704 | 0.632 | 2.696 | 7.01E-03 | 5.496 | 1.593 | 18.966 | Respiratory failure; insufficiency; arrest |
| RSP012 | C | 1.431 | 0.666 | 2.151 | 3.15E-02 | 4.184 | 1.135 | 15.421 | Respiratory failure; insufficiency; arrest |
| RSP016 | B | 0.646 | 0.265 | 2.440 | 1.47E-02 | 1.908 | 1.136 | 3.207 | Other specified and unspecified lower respiratory disease |
| RSP016 | C | 0.535 | 0.213 | 2.519 | 1.18E-02 | 1.708 | 1.126 | 2.590 | Other specified and unspecified lower respiratory disease |
| SYM002 | B | 1.305 | 0.624 | 2.092 | 3.64E-02 | 3.688 | 1.086 | 12.524 | Fever |
| SYM010 | B | 0.890 | 0.337 | 2.644 | 8.20E-03 | 2.436 | 1.259 | 4.714 | Nervous system signs and symptoms |
| SYM010 | C | 0.515 | 0.220 | 2.335 | 1.95E-02 | 1.673 | 1.086 | 2.577 | Nervous system signs and symptoms |
| SYM013 | C | 0.767 | 0.304 | 2.522 | 1.17E-02 | 2.153 | 1.186 | 3.906 | Respiratory signs and symptoms |
| SYM016 | A | 1.763 | 0.181 | 9.739 | 2.05E-22 | 5.828 | 4.088 | 8.310 | Other general signs and symptoms |
| SYM016 | B | 2.009 | 0.090 | 22.267 | 7.69E-110 | 7.456 | 6.248 | 8.899 | Other general signs and symptoms |
| SYM016 | C | 1.268 | 0.067 | 19.062 | 5.22E-81 | 3.553 | 3.119 | 4.048 | Other general signs and symptoms |
| SYM017 | B | 2.059 | 0.414 | 4.971 | 6.65E-07 | 7.835 | 3.480 | 17.640 | Abnormal findings without diagnosis |
| SYM017 | C | 1.315 | 0.289 | 4.552 | 5.32E-06 | 3.726 | 2.115 | 6.566 | Abnormal findings without diagnosis |

beta\_diff, difference in beta (regression coefficient) for the two analysis performed before and after the onset of pandemic to adjust for baseline differences between the infected and non-infected groups (see main text); boot\_se, standard error of beta\_diff; zval\_final, z-statistic in final analysis after PERR adjustment; pval\_final, corresponding p-value. Two hundred bootstraps were performed to derive the SE.

With the PERR adjustment, compared to those with no known history of COVID-19, severe COVID-19 was significantly associated with increased hazards of hospital admission due to pulmonary disorders, like pneumonia (except that caused by tuberculosis) (2.528 (1.725-3.705); p= 1.98e-06), aspiration pneumonitis (2.521 (1.099-5.786); p= 2.91e-02), respiratory failure; insufficiency; arrest (5.496 (1.593-18.966); p= 7.01e-03); and extra-pulmonary system diseases, like AMI (2.432 (1.045-5.658); p= 3.91e-02), cardiac dysrhythmias (2.219 (1.249-3.941); p= 6.57e-03), cerebral infarction (4.120 (1.989-8.535); p= 1.39e-04), peripheral and visceral vascular disease (3.331 (1.129-9.825); p= 2.93e-02), acute and unspecified renal failure (4.941 (2.918-8.364); p= 2.73e-09), UTI (3.569 (2.228-5.716); p= 1.20e-07), neurocognitive disorders (2.786 (1.160-6.689); p= 2.19e-02), transient cerebral ischemia (2.829 (1.080-7.409); p= 3.42e-02), osteomyelitis (8.640 (1.909-39.096); p= 5.12e-03), and low back pain (2.023 (1.097-3.728); p= 2.40e-02). Under PERR adjustment, we also found that higher hospitalization risks due to various infections, endocrine disorders, fever, and other general signs.

When compared with mild (non-hospitalized) COVID-19, severe COVID-19 illness (i.e. model A) were associated with higher hazards of hospitalization due to pneumonia (except that caused by tuberculosis) (2.170 (1.140-4.129); p= 1.83e-02), cardiac dysrhythmias (3.524 (1.288-9.646); p= 1.42e-02), acute and unspecified renal failure (4.025 (1.260-12.854); p= 1.87e-02), UTI (2.296 (1.142-4.614); p= 1.96e-02) and injury and intestinal infection.

Considering any (severe/mild) infection (model C), it was also observed that COVID-19 was associated with elevated hazards of hospitalization from COPD (1.635 (1.010-2.648); p= 4.55e-02), hypotension (2.069 (1.142-3.751); p= 1.66e-02), osteoarthritis (1.388 (1.101-1.750); p= 5.50e-03), and other general respiratory symptoms, in addition to most associations found under model B.

We note that the PERR adjustment tends to be conservative in general in our case. By the assumptions of PERR, the prior events should have no effect on the risk of exposure^22^; however here the history of many diseases could increase the risk of (severe) COVID-19 infection. As a result, the hazard ratio of hospitalization (for infected vs non-infected groups) will be higher than expected even in the pre-COVID-19 period, since the (severely) infected group are more likely to be hospitalized for diseases even in the pre-COVID period. This will lead to a conservative bias, as also shown in previous simulation studies^22^.

### Association with mortality after severe COVID-19

The primary results of associations with mortality after COVID-19 (without PTDM adjustment) are listed in Table 6 and Fig.4 and full results in Table S4. We found that severe COVID-19 was associated with increased all-cause mortality (HR=14.700 (13.835-15.619); p.adj<9.6e-231); significantly increased risk of mortality was also observed for mild disease, albeit with an attenuated effect size (1.237 (1.037-1.476); p.adj= 2.27e-02).

**Fig.4.**
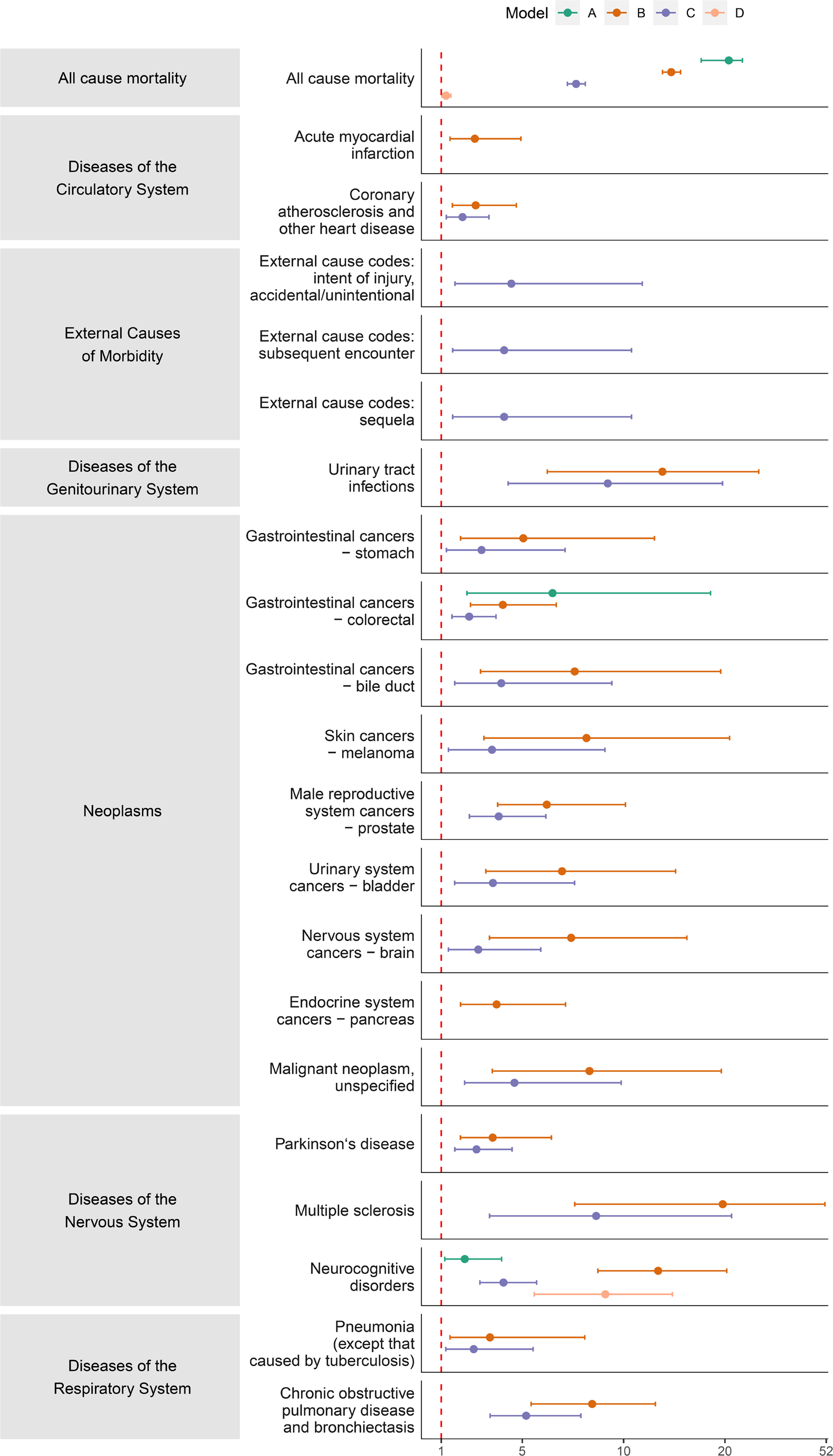
Significant association of mortality after COVID-19 (models A to D). Associations of COVID-19 with mortality from models A to D. The red dashed line stands for the line of no effect. We only present the results if the number of events>=5 for both exposed and unexposed groups.

**Table 6.** Association of mortality with severe COVID-19, without PTDM adjustment.

| Outcome | coef | exp(coef) | se(coef) | z | Pr(>\|z\|) | p.adj | 2.5.% | 97.5.% | Model | Analysis | CCSR Category Description |
| --- | --- | --- | --- | --- | --- | --- | --- | --- | --- | --- | --- |
| All-cause Mortality | 3.055 | 21.229 | 0.094 | 32.536 | 3.27E-232 | **9.60E-231** | 17.661 | 25.520 | A | All-cause mortality |  |
| All-cause Mortality | 2.688 | 14.700 | 0.031 | 86.872 | 0.00E+00 | **0.00E+00** | 13.835 | 15.619 | B | All-cause mortality |  |
| All-cause Mortality | 2.035 | 7.655 | 0.029 | 69.261 | 0.00E+00 | **0.00E+00** | 7.226 | 8.109 | C | All-cause mortality |  |
| All-cause Mortality | 0.213 | 1.237 | 0.090 | 2.360 | 1.83E-02 | **2.27E-02** | 1.037 | 1.476 | D | All-cause mortality |  |
| NEO015 | 1.871 | 6.493 | 0.537 | 3.487 | 4.89E-04 | **8.30E-04** | 2.269 | 18.583 | A | Any death | Gastrointestinal cancers - colorectal |
| NVS011 | 0.771 | 2.162 | 0.311 | 2.475 | 1.33E-02 | **1.70E-02** | 1.174 | 3.980 | A | Any death | Neurocognitive disorders |
| CIR009 | 0.978 | 2.658 | 0.315 | 3.100 | 1.94E-03 | **2.89E-03** | 1.432 | 4.931 | B | Any death | Acute myocardial infarction |
| CIR011 | 0.993 | 2.700 | 0.284 | 3.504 | 4.59E-04 | **8.25E-04** | 1.549 | 4.707 | B | Any death | Coronary atherosclerosis and other heart disease |
| GEN004 | 2.627 | 13.832 | 0.407 | 6.461 | 1.04E-10 | **5.70E-10** | 6.235 | 30.687 | B | Any death | Urinary tract infections |
| NEO013 | 1.619 | 5.050 | 0.484 | 3.344 | 8.25E-04 | **1.34E-03** | 1.955 | 13.048 | B | Any death | Gastrointestinal cancers - stomach |
| NEO015 | 1.396 | 4.040 | 0.256 | 5.448 | 5.08E-08 | **1.94E-07** | 2.445 | 6.676 | B | Any death | Gastrointestinal cancers - colorectal |
| NEO018 | 2.026 | 7.587 | 0.484 | 4.189 | 2.80E-05 | **6.16E-05** | 2.940 | 19.582 | B | Any death | Gastrointestinal cancers - bile duct |
| NEO025 | 2.100 | 8.168 | 0.493 | 4.256 | 2.08E-05 | **4.82E-05** | 3.105 | 21.487 | B | Any death | Skin cancers - melanoma |
| NEO039 | 1.826 | 6.208 | 0.252 | 7.233 | 4.72E-13 | **3.19E-12** | 3.785 | 10.181 | B | Any death | Male reproductive system cancers - prostate |
| NEO043 | 1.940 | 6.962 | 0.396 | 4.898 | 9.68E-07 | **2.94E-06** | 3.203 | 15.133 | B | Any death | Urinary system cancers - bladder |
| NEO048 | 2.003 | 7.411 | 0.401 | 5.001 | 5.71E-07 | **1.93E-06** | 3.380 | 16.249 | B | Any death | Nervous system cancers - brain |
| NEO051 | 1.317 | 3.732 | 0.331 | 3.984 | 6.79E-05 | **1.39E-04** | 1.952 | 7.134 | B | Any death | Endocrine system cancers - pancreas |
| NEO071 | 2.118 | 8.314 | 0.439 | 4.828 | 1.38E-06 | **3.92E-06** | 3.519 | 19.646 | B | Any death | Malignant neoplasm, unspecified |
| NVS004 | 1.264 | 3.540 | 0.305 | 4.143 | 3.43E-05 | **7.37E-05** | 1.947 | 6.439 | B | Any death | Parkinson`s disease |
| NVS005 | 2.985 | 19.784 | 0.489 | 6.105 | 1.03E-09 | **4.76E-09** | 7.588 | 51.582 | B | Any death | Multiple sclerosis |
| NVS011 | 2.596 | 13.406 | 0.219 | 11.858 | 1.95E-32 | **4.28E-31** | 8.729 | 20.588 | B | Any death | Neurocognitive disorders |
| RSP002 | 1.225 | 3.404 | 0.441 | 2.777 | 5.49E-03 | **7.55E-03** | 1.434 | 8.083 | B | Any death | Pneumonia (except that caused by tuberculosis) |
| RSP008 | 2.134 | 8.448 | 0.225 | 9.479 | 2.57E-21 | **3.23E-20** | 5.434 | 13.134 | B | Any death | Chronic obstructive pulmonary disease and bronchiectasis |
| CIR011 | 0.718 | 2.050 | 0.251 | 2.855 | 4.31E-03 | **6.02E-03** | 1.252 | 3.355 | C | Any death | Coronary atherosclerosis and other heart disease |
| EXT020 | 1.495 | 4.459 | 0.498 | 2.999 | 2.71E-03 | **3.97E-03** | 1.679 | 11.844 | C | Any death | External cause codes: intent of injury, accidental/unintentional |
| EXT029 | 1.412 | 4.105 | 0.493 | 2.866 | 4.15E-03 | **5.90E-03** | 1.563 | 10.781 | C | Any death | External cause codes: subsequent encounter |
| EXT030 | 1.412 | 4.105 | 0.493 | 2.866 | 4.15E-03 | **5.90E-03** | 1.563 | 10.781 | C | Any death | External cause codes: sequela |
| GEN004 | 2.221 | 9.217 | 0.389 | 5.706 | 1.15E-08 | **4.83E-08** | 4.298 | 19.766 | C | Any death | Urinary tract infections |
| NEO013 | 1.095 | 2.990 | 0.442 | 2.477 | 1.33E-02 | **1.70E-02** | 1.257 | 7.113 | C | Any death | Gastrointestinal cancers - stomach |
| NEO015 | 0.867 | 2.380 | 0.226 | 3.840 | 1.23E-04 | **2.35E-04** | 1.529 | 3.704 | C | Any death | Gastrointestinal cancers - colorectal |
| NEO018 | 1.377 | 3.963 | 0.442 | 3.117 | 1.83E-03 | **2.77E-03** | 1.667 | 9.421 | C | Any death | Gastrointestinal cancers - bile duct |
| NEO025 | 1.254 | 3.504 | 0.486 | 2.582 | 9.82E-03 | **1.31E-02** | 1.353 | 9.078 | C | Any death | Skin cancers - melanoma |
| NEO039 | 1.345 | 3.837 | 0.242 | 5.560 | 2.70E-08 | **1.08E-07** | 2.389 | 6.165 | C | Any death | Male reproductive system cancers - prostate |
| NEO043 | 1.268 | 3.552 | 0.387 | 3.277 | 1.05E-03 | **1.68E-03** | 1.664 | 7.582 | C | Any death | Urinary system cancers - bladder |
| NEO048 | 1.039 | 2.827 | 0.376 | 2.762 | 5.75E-03 | **7.79E-03** | 1.352 | 5.912 | C | Any death | Nervous system cancers - brain |
| NEO071 | 1.529 | 4.614 | 0.389 | 3.935 | 8.32E-05 | **1.66E-04** | 2.154 | 9.881 | C | Any death | Malignant neoplasm, unspecified |
| NVS004 | 1.008 | 2.741 | 0.253 | 3.991 | 6.57E-05 | **1.38E-04** | 1.671 | 4.497 | C | Any death | Parkinson`s disease |
| NVS005 | 2.157 | 8.645 | 0.479 | 4.506 | 6.61E-06 | **1.62E-05** | 3.383 | 22.092 | C | Any death | Multiple sclerosis |
| NVS011 | 1.404 | 4.072 | 0.172 | 8.159 | 3.37E-16 | **2.96E-15** | 2.906 | 5.706 | C | Any death | Neurocognitive disorders |
| RSP002 | 0.957 | 2.603 | 0.385 | 2.485 | 1.29E-02 | **1.70E-02** | 1.224 | 5.535 | C | Any death | Pneumonia (except that caused by tuberculosis) |
| RSP008 | 1.647 | 5.190 | 0.214 | 7.706 | 1.30E-14 | **1.04E-13** | 3.414 | 7.890 | C | Any death | Chronic obstructive pulmonary disease and bronchiectasis |
| NVS011 | 2.208 | 9.100 | 0.249 | 8.880 | 6.66E-19 | **7.33E-18** | 5.590 | 14.816 | D | Any death | Neurocognitive disorders |

Coef, regression coefficient from Cox model. HR, hazard ratio; se, standard error; p.adj, FDR-adjusted p-value; Lower and upper CI represent the lower and upper 95% confidence interval of HR.

Considering cause-specific mortality (without considering the past history of disease), severe infection was associated with mortality due to pneumonia (except that caused by tuberculosis) (3.404 (1.434-8.083); p.adj= 7.55e-03), COPD (8.448 (5.434-13.134); p.adj= 3.23e-20), AMI (2.658 (1.432-4.931); p.adj= 2.89e-03), coronary atherosclerosis and other heart disease (2.700 (1.549-4.707); p.adj= 8.25e-04), Parkinson’s disease (3.540 (1.947-6.439); p.adj= 7.37e-05), multiple sclerosis (19.784 (7.588-51.582); p.adj= 4.76e-09), neurocognitive disorders (13.406 (8.729-20.588); p.adj= 4.28e-31), and death due to several cancers (see Table 6).

Associations with mortality after any infection (model C) are largely similar. When we only considered fatalities from those with a history of the disease, most associations were non-significant, likely due to the small number of events and lower statistical power. Associations with mortality with PTDM adjustment is similar to those without PTDM adjustment (Table S4 and Fig.S3).

### Analysis restricted to sequelae 15 or 30 days post infection

In an additional sensitivity analysis, we restrict outcomes that occur 15 or 30 days after the date of infection (Table S7). The overall patterns of sequelae were similar.

### Analysis restricted to the pre-vaccination period

Association of all hospitalization and mortality after COVID-19 before the launch of vaccination (i.e. prior to 8 Dec 2020) are presented in Tables S5 (also see Fig.S4) and S6 (also see Fig.S5) respectively. The spectrum of sequelae remained similar. As for mortality, increased hazards of all-cause mortality and mortality due to specific common causes such as coronary atherosclerosis and other heart disease, acute hemorrhagic cerebrovascular disease, COPD, and other cancers were also observed when the analysis was limited to the ‘pre-vaccination’ period.

When comparing the HR for various sequelae in the pre-vaccination vs the whole follow-up (FU) periods, the estimated HR for hospitalization in the *pre*-vaccination period was generally higher (Table S6b). For example under model B, out of the 65 disorders included in this analysis, 55 showed higher HRs for hospitalization in the pre-vaccination era (binomial p, one-tailed= 5.88E-09); similar result was observed for model C (62 out of 73 disorders showing higher HRs in the pre-vaccination period, p=4.55E-10). For individual disease categories, for example, we found higher HRs in the pre-vaccination period for circulatory disorders (8 out of 9) and respiratory disorders (5 out of 5) and several other categories (model B; please refer to Table S6b for details). Note that we restricted to disorders with at least 5 events in each group such that a meaningful comparison can be made. For ‘other general signs and symptoms’, the HR in the pre-vaccination period was substantially higher when compared to the estimate based on the whole FU period, with non-overlapping CIs. In addition, we observed that for all-cause mortality (under model A/B/C), HR of the pre-vaccination period were substantially higher than the estimate from the whole FU period, with non-overlapping CIs and significant p-values (p=5.62E-03, 1.3eE-21 and 1.40E-42 under model A, B and C respectively). We computed standard error (SE) of the difference in coefficients using non-parametric bootstrap. We note however that the HR estimates of the pre-vaccination period were generally less precise, due to smaller number of events.

### Associations results stratified by vaccination status

The associations with sequelae stratified by vaccination status are presented in Table S8 and Fig.S6. We will focus on the comparison between subjects with vaccination before (and to a lesser extent, after) the infection against those infected but have not received any vaccination. A few results were nominally significant. For the analysis on breakthrough infections vs infection without vaccination, we observed lower risks of hospitalization from pneumonia, unspecified lower respiratory diseases and other general signs/symptoms from the former group. We observed increased rates of hospitalization from a few non-specific causes, including low back pain, non-specific chest pain and cataracts. However, such results may not reflect genuine causal relationships as the exact reason for admission is unknown (e.g. admission could be for an elective operation). The most significant difference was observed for general signs and symptoms (p=1.83e-15; lower HR for breakthrough infections).

We then compared those vaccinated after infection against those infected and unvaccinated. The former group had lower hazards of hospitalization from injuries, pneumonia, bacterial infections, septicaemia and other general signs and symptoms, under the original model without PTDM adjustment.

Nevertheless, for subjects who received vaccinations after being infected, we only consider events occurring after vaccination, when comparing with infected but unvaccinated subjects (for whom we will consider all events after infection). As such, the PTDM adjustment may produce more reasonable results. No results were nominally significant under PTDM except for urinary tract infections which was marginally significant.

### Associations results stratified by follow-up period (time lapsed after infection)

The proportional hazards assumption was violated in some analyses (considering all hospitalization as outcome) (Table S9). We performed time-stratified and time-transformation analysis to estimate time-dependent hazard ratios. As for the analysis stratified by follow-up period, we observed larger effects from COVID-19 infection and more statistically significant associations when restricting analysis of outcome to the first 3 months, and in general the effects of infection attenuated over time (Table S10 and Fig S7). Nevertheless, it is noteworthy that even after 6 months, infections and severe infections in particular, were still associated with elevated hospitalization risks from numerous conditions. These included, for example, aspiration pneumonitis, acute renal failure, injuries and fractures, and various infections such as urinary tract infections, septicaemia, pneumonia and other bacterial infections (Table S10). The results of tests for proportional hazards for time-stratified analysis are shown in Table S11. Analyses using time-transformation are presented in Table S12. For the majority of outcomes, the baseline risk was the highest, and the risk decreased over time. However, the estimated time taken (assuming a linear reduction in risk on the scale of log(*t*)) for the adverse effects of infection to wane to zero was generally over 6 months. As an example, the median time taken for the deleterious effects of severe infection (i.e. model B) to wane to zero was 403.8 days. Results on mortality using time-split and time-transformation analyses are presented in Table S13.

#### Patient and public involvement

Patients were not involved in the conception, design or implementation of the study. No patients were involved in data interpretation or preparation of this paper. This work was conducted to address research concerns in view of the escalating COVID-19 pandemic, and we believe is of public health interest, although no patients/public was directly involved due to the need to complete the study in a timely manner.

## Discussion

### Overview

Overall, we found that COVID-19, especially severe disease, were associated with increased risks of hospitalization and/or mortality due to pulmonary, cardiovascular, digestive, neurological, genitourinary and musculoskeletal disorders in the post-infection period. These results were largely consistent and robust to multiple sensitivity analysis, and after PERR and PTDM adjustment.

### Interpretation of findings in the context of previous studies

We noted that a recent study by Al-Aly et al. has characterized the post-acute sequelae of COVID-19 in US veterans^9^ for a range of outcomes, however the study was on veterans only which may affect its generalizability to the general population and females. The focus of Al-Aly et al. was also somewhat different from ours, as they focused on incident (new) diagnoses post-infection, while we studied *hospitalization* due to new or recurrent diseases. As such, a person with history of a disorder (e.g. stroke) who was admitted for the same disease post-infection will be considered by our study but not in Al-Aly et al. We also studied mortality due to all disorders covering all systems.

In this study we utilized a large prospective cohort of UKBB to delineate the septum of hospital admission and mortality cause up to 20 months after COVID-19. We have conducted a very comprehensive set of analyses, which includes all hospitalization and mortality from diseases covering all systems, as well as stratified analysis on new-onset and recurrent diseases. We also performed other sensitivity analyses such as PTDM and PEER adjustments. The PEER adjustment for example can help to reduce residual confounding and evaluate causal relationships between COVID-19 and possible sequelae, which has not been employed in the above study^9^.

A number of studies have been performed on the longer-term sequelae of COVID-19 (mostly focused on specific organ systems), and have revealed elevated risks for a variety of disorders, which corroborate with our findings in general. For example, impaired lung function and respiratory conditions are among the most commonly reported sequelae of COVID-19^3,4^. Two previous studies on US veterans^8,9^ have revealed that COVID-19 was associated with a wide range of cardiovascular disorders including dysrhythmias, inflammatory and ischemic heart diseases, thrombotic disorders, heart failure, cerebrovascular conditions and others. The current study also revealed a similar spectrum of cardiovascular in a general population cohort based on an independent sample. Cerebrovascular disorders were also commonly reported as complications of COVID-19, as reviewed elsewhere^25,26^. Also, renal involvement is not uncommon during the acute phase of infection, and subclinical inflammation may persist for many months^27,28^. Deterioration of renal function following acute infection has been reported^11^. Although the mechanisms remain poorly understood, neurocognitive impairment was observed for patients after recovery from COVID-19^29-31^. In a large-scale retrospective cohort study^32^, it was reported that the incidence of most neurological and psychiatric disorders were raised after COVID-19, compared to those with influenza or other respiratory tract infections, especially for the most severe COVID-19 cases requiring intensive care support.

We also observed that COVID-19 patients were more likely to be admitted for other bacterial or viral infections post-infection. Elevated risk of hospitalization was not only observed for pneumonia, but also other infective conditions such as septicemia, peritonitis and intra-abdominal abscess, intestinal infection, urinary tract infections (UTI), osteomyelitis, and other bacterial infections. The exact mechanisms remain unclear, however for post-influenza infections, it has been suggested that secondary infections could be due to disruption of airway epithelium owing to immune-mediated damage and dysfunctional innate immune responses^33,34^. Since corticosteroids are often prescribed for severe COVID-19 infections, it may be worthwhile to investigate whether steroid use is associated with increased risks of other infections^35^.

Another interesting finding is that we observed increased risk of hospitalization from several musculoskeletal problems, especially fractures. Increased risks of fall may be one important contributing factor, which in turn could be due to myriad reasons such as malaise and fatigue, musculoskeletal pain, muscle weakness, neurocognitive impairment, poor general well-being and the increased incidence of other disorders that may increase fall risk post-infection^9,36^. Another contributing factor may be increased risks of osteoporosis due to COVID-19 itself^37^ and/or its treatment (e.g. steroids), but further clinical studies are required. We also noted increased risks of hospitalization from several neoplasms, which may be due to, for example, deconditioning and increased risk of secondary infections after the acute COVID-19 infection, given that it is unlikely that COVID-19 would affect the incidence of neoplasms.

While this is an observational study, it is interesting to note that Mendelian randomization studies, which are less prone to confounding, have also shown that COVID-19 may be a causal risk factor for various cardiometabolic and neuropsychiatric disorders^38,39^.

The mechanisms underlying COVID-19 sequelae are still largely unknown, and different symptoms/disorders may also involve different pathophysiology. It has been postulated that, for example, persistent reservoirs of the virus, on-going inflammation, dysregulated immune response and re-activation of pathogens may all contribute to the long-term sequelae^40,41^.

Whether vaccination may ameliorate the risk of COVID-19 sequelae is another important question. Interestingly, we observed preliminary evidence that the risks of sequelae were in general higher in the pre-vaccination period, and there was a higher-than-expected proportion of diseases which showed larger HR in the pre-vaccination era. For all-cause mortality, HR was also significantly higher before the introduction of COVID-19 vaccines. Moreover, we observed that for some sequelae (e.g. non-COVID-19 pneumonia, septicaemia, bacterial infections, other general signs/symptoms), subjects who are vaccinated before or after infection had lower risks than infected patients who are not vaccinated.

However, this study is not primarily designed to study COVID-19 vaccinations, and the results require replications in future studies. This study is observational in nature, and the sample size may still be inadequate to capture uncommon events. We cannot exclude the possibility that apparently higher risks of sequelae in the pre-vaccination era could also be due to other time-varying factors (e.g. different circulating variants or treatment strategies). However, our study period was before the emergence of Omicron, and other variants during the post-vaccination period, such as Alpha and Delta, are likely to be of at least similar or higher pathogenicity than the original strain of virus^42^. Anti-viral medications such as Paxlovid and molnupiravir were not yet available in the study period. In summary, it is encouraging to observe preliminary evidence that vaccination may reduce the risk of sequelae, but the findings should be regarded as tentative before more evidence are available.

### Clinical implications

Our findings may have important clinical implications. We observed that COVID-19, especially severe infections requiring hospitalization, is associated with a wide variety of pulmonary and extra-pulmonary sequelae. Alarmingly, elevated risk of admission and mortality was observed for multiple disorders spanning many organ systems. Patients who have recovered from especially severe COVID-19 may therefore require appropriate follow-up, monitoring and risk assessment for different complications. It remains to be investigated what types of interventions may help to reduce the risk of sequelae.

From a public health perspective, when determining policies related to the pandemic (such as lifting restrictions), it may be advisable to consider the total burden caused by COVID-19 that includes its possible complications, and the disabilities and mortalities due to these complications. Our findings also call for allocation of resources to prevent and treat the relevant complications.

Another alarming finding is that even mild (non-hospitalized) COVID-19 may be associated with modestly increased risk of all-cause mortality (HR=1.237, 95% CI 1.037-1.476) and mortality from neurocognitive disorders, as well as hospital admission from a few disorders such as aspiration pneumonitis, musculoskeletal pain and other general signs/symptoms. Such findings are in line with previous studies^25,26^ which also showed elevated risks of cardiovascular and other sequelae even for mild COVID-19 cases. In Al-Aly et al.^9^, it was reported that mild (non-hospitalized) COVID-19 had elevated mortality risks (HR=1.59 (1.46-1.73)) beyond the first 30 days of disease, which corroborates with our findings. The difference in the effect size could be due to heterogeneity of the two samples^43^, different gender proportions, and lengths of follow-up etc.

Given the huge number of people infected by COVID-19, even a modest rise in the hazard for mortality would imply a large (absolute) number of fatalities. Similarly, the absolute burden and disabilities caused by hospitalizations related to mild infections could be large. This suggests containment of infection is still important together with efforts to reduce the number of severe cases.

### Strengths and limitations

The major strength of this study included large sample size of the UK Biobank with detailed health records and sufficient period of follow-up. This study is very comprehensive and covers most clinically relevant categories of diagnosis. In addition to any hospitalization/mortality, we also performed additional analysis on hospitalization with/without relevant history of disease (i.e. new-onset and recurrent diseases). Apart from these, we also made use of advanced statistical methods to evaluate whether our findings are consistent to different modeling strategies. For example, the PERR adjustment was employed to minimize unmeasured confounding.

There are several limitations of this study. Firstly, this is an observational study. Although we have included many known confounders in the model and used other methods (e.g. PERR) to minimize residual confounding, confounding effects cannot be excluded completely and causality cannot be concluded. However, many results remained significant after PERR adjustment which provides further support to our findings. Secondly, subclinical disorders may be underestimated and some disorders may be too rare to be included for further analysis. In addition, due to the relatively small number of events for some disorders, absence of significant associations can also be due to inadequate statistical power. Small number of events (e.g. mortality from specific disorders) may also lead to relatively imprecise effect estimates and wide confidence intervals. We also note that mild or asymptomatic infections may not be detected and considered as having no history of COVID-19; however this is likely to result in a conservative bias. Variant information is not known and thus cannot be analyzed, although we expect the Alpha and Delta variants are likely the predominant variants. Further studies are required to investigate the sequelae from other variants, such as Omicron. Finally, UKBB participants are likely healthier and of higher socioeconomic status^44^, and generalizability of current findings to other populations (e.g. other ethnicities and those aged<50) may require further studies.

## Conclusions

In conclusion, this study reveals increased risk of hospitalization and mortality from a wide variety of pulmonary and extra-pulmonary diseases after COVID-19, especially for those with severe disease. On the other hand, we also observed increased all-cause mortality and hospitalization risk from several disorders after mild infections. The findings may have important clinical and public health implications given the huge number of people infected by COVID-19. Proper monitoring and assessments for the risk of sequelae may be warranted for patients recovered from severe COVID-19 in particular. Further studies are required to replicate the current findings and to investigate the mechanisms underlying the sequelae, and interventions for prevention and treatment.

## Supporting information

List of supp info

## Data Availability

The UK Biobank data is available to all registered researchers upon application. All other data produced in the present work are contained in the manuscript.

## Author Contributions

Conception and design: HCS (lead), with input from YX. Study supervision: HCS. Funding acquisition: HCS. Methodology: HCS (lead), YX. Data curation: YX (lead), RZ, JQ. Data analysis: YX (lead), RZ, JQ. Data interpretation: HCS, YX, RZ, JQ. Preparation of first draft of manuscript: YX, HCS, with input from RZ and JQ.

## Supplementary Information

All supplementary Tables and notes are available at the journal’s website and at https://drive.google.com/drive/folders/1E2hsN0E7wwuTa7mqxLHmAizz9j_Cxc?usp=sharing

## Acknowledgements

This work was supported partially a National Natural Science Foundation China (NSFC) grant (81971706) and the Lo Kwee Seong Biomedical Research Fund from The Chinese University of Hong Kong and the KIZ-CUHK Joint Laboratory of Bioresources and Molecular Research of Common Diseases, Kunming Institute of Zoology and The Chinese University of Hong Kong, China. We thank Prof. Pak Sham for support on data access. We also thank Prof. Stephen Tsui and Prof. Simon Lui for useful discussions.

