## Supplementary material for "Association of COVID-19 with risks of hospitalization and mortality from other disorders post-infection: A study of the UK Biobank": List of supp info

All supplementary Tables and notes are available at the journal’s website and at <https://drive.google.com/drive/folders/1E2hsN0E7wwuT__a7mqxLHmAizz9j_Cxc?usp=sharing> .

**Supplementary Tables**

Table S1	Full list of covariates and distribution of covariates

Table S2	Missing rate and out-of-bag prediction error from imputation

Table S3	Association of COVID-19 with hospitalization due to various diseases, models A to D (full table containing analysis on all hospitalizations, new-onset and recurrent diseases and analysis with or without PTDM)

Table S4	Association of COVID-19 with mortality due to various diseases, models A to D (full table containing analysis on all hospitalizations, new-onset and recurrent diseases and analysis with or without PTDM)

Table S5 Association of COVID-19 with hospitalization due to various diseases, models A to D, restricted to the pre-vaccination period (before 8 Dec 2020)

Table S6	Association of COVID-19 with mortality due to various diseases, models A to D, restricted to the pre-vaccination period (before 8 Dec 2020)

[Note to Table S6: Table S6a shows all association test results; Table S6b shows binomial test of the proportion of times that the HR from the pre-vaccination period is higher than that from the whole follow-up period]

Table S7	Association of COVID-19 and hospitalization/mortality due to various diseases, restricted to events that occur 15 or 30 days after being tested positive for COVID-19

Table S8	Association of COVID-19 with hospitalization due to various diseases, as stratified by vaccination status (e.g. before or after infection)

[Note to Table S8: We performed further analyses and compared hospitalization rates for subjects who have received at least one dose of vaccine *before* infection, those who have received vaccination *after* the infection and those who are infected but not vaccinated; full results are presented in Table S8a, and Table S8b highlighted findings which focuses on comparison against infected subjects who are un-vaccinated]

Table S9	Tests of proportional hazards assumption (for association tests of all hospitalizations)

Table S10	The association of COVID-19 infection and risk of subsequent hospitalization within different time periods

Table S11	The tests of proportional hazards assumption for time-split analysis with any hospitalization

Table S12	The association of COVID-19 infection and risk of subsequent hospitalizations using time- transformation analysis (see the main text and table legends for details)

Table S13 Associations with COVID-19 and subsequent mortality using time splits and time transformation

**Supplementary Figures:**

Fig.S1	Association of COVID-19 with hospitalization due to various diseases, models A to D (containing analysis on all hospitalizations, new-onset and recurrent diseases and analysis without PTDM)

Legend: associations of COVID-19 with hospitalization without PTDM adjustment. The red dashed line stands for the line of no effect (hazard ratio=1). Effect estimates lying on the right of the red dashed line indicates elevated hazards (HR>1). X-axis indicated the HR (hazard ratio) of hospitalization post COVID-19. Y-axis showed each specific diagnoses category. Confidence intervals are also shown. For

Fig.S2	Association of COVID-19 with hospitalization due to various diseases, models B to D  (full table containing analysis on all hospitalizations and analysis with and without PTDM)

Fig.S3	Association of COVID-19 with mortality due to various diseases, models B to D, with and without PTDM adjustment

Fig.S4	 Association of COVID-19 with hospitalization due to various diseases, models A to D, restricted to pre-vaccination period  (before 8 Dec 2020), plot with whole period.

Fig.S5	Association of COVID-19 with mortality due to various diseases, models A to D, restricted to pre-vaccination period (before 8 Dec 2020) , plot with whole period

Fig.S6 Association of COVID-19 with hospitalization due to various diseases, as stratified by vaccination status.

Fig.S7 Association of COVID-19 infection and risk of subsequent hospitalization within different time periods

(please refer to the legend of Fig S1 for other figures S2-7)
